# Interleukin-6 signaling effects on ischemic stroke and other cardiovascular outcomes: a Mendelian Randomization study

**DOI:** 10.1101/19007682

**Authors:** Marios K. Georgakis, Rainer Malik, Dipender Gill, Nora Franceschini, Cathie L. M. Sudlow, INVENT Consortium, CHARGE Inflammation Working Group, Martin Dichgans

## Abstract

**Background:** Studies in humans and experimental models highlight a role of interleukin-6 (IL-6) in cardiovascular disease. Indirect evidence suggests that inhibition of IL-6 signaling could lower risk of coronary artery disease. However, whether such an approach would be effective for ischemic stroke and other cardiovascular outcomes remains unknown.

**Methods:** In a genome-wide association study (GWAS) of 204,402 European individuals, we identified genetic proxies for downregulated IL-6 signaling as genetic variants in the IL-6 receptor (*IL6R*) locus that were associated with lower C-reactive protein (CRP) levels, a downstream effector of IL-6 signaling. We then applied two-sample Mendelian randomization (MR) to explore associations with ischemic stroke and its major subtypes (large artery stroke, cardioembolic stroke, small vessel stroke) in the MEGASTROKE dataset (34,217 cases and 404,630 controls), with coronary artery disease in the CARDIoGRAMplusC4D dataset (60,801 cases and 123,504 control), and with other cardiovascular outcomes in the UK Biobank (up to 321,406 individuals) and in phenotype-specific GWAS datasets. All effect estimates were scaled to the CRP-decreasing effects of tocilizumab, a monoclonal antibody targeting IL-6R.

**Results:** We identified 7 genetic variants as proxies for downregulated IL-6 signaling, which showed effects on upstream regulators (IL-6 and soluble IL-6R levels) and downstream effectors (CRP and fibrinogen levels) of the pathway that were consistent with pharmacological blockade of IL-6R. In MR, proxies for downregulated IL-6 signaling were associated with lower risk of ischemic stroke (Odds Ratio [OR]: 0.89, 95%CI: 0.82-0.97) and coronary artery disease (OR: 0.84, 95%CI: 0.77-0.90). Focusing on ischemic stroke subtypes, we found significant associations with risk of large artery (OR: 0.76, 95%CI: 0.62-0.93) and small vessel stroke (OR: 0.71, 95%CI: 0.59-0.86), but not cardioembolic stroke (OR: 0.95, 95%CI: 0.74-1.22). Proxies for IL-6 signaling inhibition were further associated with a lower risk of myocardial infarction, aortic aneurysm, atrial fibrillation and carotid plaque.

**Conclusions:** We provide evidence for a causal effect of IL-6 signaling on ischemic stroke, particularly large artery and small vessel stroke, and a range of other cardiovascular outcomes. IL-6R blockade might represent a valid therapeutic target for lowering cardiovascular risk and should thus be investigated in clinical trials.

**CLINICAL PERSPECTIVE:** *What is new:*

- We identified genetic proxies for downregulated IL-6 signaling that had effects on upstream and downstream regulators of the IL-6 signaling pathway consistent with those of pharmacological IL-6R blockade
- Genetically downregulated IL-6 signaling was associated with a lower risk of ischemic stroke, and in particular large artery and small vessel stroke
- Similar associations were obtained for a broad range of other cardiovascular outcomes

*What are the clinical implications:*

- Inhibition of IL-6 signaling is a promising therapeutic target for lowering risk of stroke and other cardiovascular outcomes and should be further investigated in clinical trials

## INTRODUCTION

Stroke is the leading cause of adult disability and the second most common cause of mortality worldwide^1, 2^ with an increasing burden on global health.^3, 4^ Inflammation is involved in the pathogenesis of ischemic stroke, as has specifically been demonstrated for large artery atherosclerotic stroke.^5, 6^ Cytokines regulate inflammatory responses^5^ and could thus serve as targets for cardiovascular disease prevention.^7^ In the recent Canakinumab Anti-Inflammatory Thrombosis Outcomes Study (CANTOS), treatment with an interleukin-1β (IL-1β) antagonist reduced cardiovascular event rates in patients with a history of myocardial infarction.^8^ However, whether interfering with other cytokines would likewise offer benefit remains largely unknown. Also, there are few data on stroke and other cardiovascular outcomes beyond coronary artery disease.^9-11^

Interleukin-6 (IL-6), a key regulator of the inflammatory cascade, acts by binding to either its membrane-bound or soluble receptor (IL-6R) and induces proinflammatory downstream effects including increases in the levels of C-reactive protein (CRP).^12, 13^ IL-6 has been implicated in the pathogenesis of multiple inflammatory diseases and inhibitors of IL-6R are used for the treatment of rheumatoid arthritis,^14^ inflammatory bowel disease,^15^ and other autoimmune disorders.^16^ Downregulation of IL-6 signaling has further been proposed as a potential strategy for lowering cardiovascular risk.^11, 13^ IL-6 levels have consistently been associated with risk of coronary artery disease in cohort studies.^17, 18^ Mendelian randomization (MR) studies further showed that a variant in the gene encoding IL-6R with effects resembling pharmacological IL-6R inhibition is associated with a lower risk of coronary artery disease.^19, 20^ Finally, secondary analyses from CANTOS demonstrated that the magnitude of the therapeutic benefit of IL-1β targeting was associated with the reduction of circulating IL-6 levels^11, 21^ and that even after IL-1β inhibition, the residual cardiovascular risk was proportional to the post-treatment IL-6 levels.^22^ These results provide indirect clinical evidence that interfering with IL-6 signaling might lower cardiovascular risk and suggest that an approach directly targeting IL-6 signaling could offer additional benefit for cardiovascular prevention beyond IL-1β inhibition.

The effects of IL-6 signaling on risk of ischemic stroke remain largely unknown. While population-based cohort studies have found that circulating IL-6 levels are associated with a higher risk of ischemic stroke,^23, 24^ these associations preclude conclusions about causal relationships because of possible confounding and reverse causation bias.^25^ Also, there are no data on etiological stroke subtypes and other cardiovascular outcomes beyond coronary artery disease. Developing meaningful strategies for the prevention of ischemic stroke and cardiovascular disease in general would require defining these relationships.^26^

By using genetic variants as proxies for a trait of interest, MR overcomes key limitations of observational studies such as confounding and reverse causation and allows for investigation of causal effects on outcomes.^27, 28^ MR further allows for prediction of the effects of pharmacological interventions by using variants located close to genes encoding candidate drug targets.^29, 30^ Hence, MR has become a powerful strategy to prioritize interventions for exploration in clinical trials.^28^

Here, leveraging data from large genome-wide association studies (GWASs)^31-33^ and applying MR analyses, we aimed to: (i) identify genetic proxies for downregulated IL-6 signaling on the basis of their effects on CRP levels, a well-established IL-6 signaling downstream effector,^13, 20, 34^ (ii) validate their utility by comparing the consistency of their effects on upstream regulators and downstream effectors of the IL-6 signaling pathway with the effects of pharmacological IL-6R inhibition, as derived from clinical trials, (iii) explore associations of genetic predisposition to downregulated IL-6 signaling with the risk of ischemic stroke and coronary artery disease, (iv) examine associations with major etiological subtypes of ischemic stroke (large artery, cardioembolic, and small vessel stroke), and (v) examine associations with a broad range of other cardiovascular phenotypes. To derive clinically meaningful effect sizes that would be comparable to those derived from potential future clinical trials, we weighted our instruments based on the CRP-decreasing effects of tocilizumab, a monoclonal antibody targeting IL-6R.

## METHODS

### Selection of genetic proxies for IL-6 signaling and validation of the instruments

The data sources for this study are described in **Table 1**. To identify instruments for genetic predisposition to downregulated IL-6 signaling, we selected variants within or near the *IL6R* gene, which encodes the receptor of IL-6. Specifically, we selected single-nucleotide polymorphisms (SNPs) in the *IL6R* gene or a region of 300 kB upstream or downstream from the *IL6R* gene (GRCh37/hg19 coordinates: chr1:154,077,669-154,741,926; **Supplementary Figure 1**) that were associated with circulating CRP levels. We selected and weighted genetic instruments for genetic predisposition to IL-6 signaling on the basis of their associations with CRP levels, because elevated CRP levels are a well-described downstream effect of IL-6 signaling (**Figure 1**).^13, 20, 34^ Genetic association estimates with circulating CRP levels were obtained from a GWAS of 204,402 individuals of European ancestry drawn from the Cohorts for Heart and Aging Research in Genomic Epidemiology (CHARGE) Inflammation Working Group.^31^ We selected variants that were associated with circulating CRP levels at genome-wide significance (*p*<5×10^−8^) and clumped these variants to a linkage disequilibrium (LD) threshold of *r*^*2*^ < 0.1 according to the European reference panel of the 1000 Genomes project.^35^ We estimated the variance in CRP levels explained by each of the SNPs by calculating the *R*^*2*^,^36^ and the strength of the instruments by calculating the F-statistic.^37^

**Table 1.** Data sources that were used in the analyses for the current study.

| Phenotype | Source | N (Total or Cases/Controls) | Imputation reference panel | Ancestry | Adjustments |
| --- | --- | --- | --- | --- | --- |
| CRP levels | CHARGE Inflammation Working Group<sup>31</sup> | 204,402 | HapMap | European | age, sex, population structure |
| IL-6 levels | YFS/FINRISK studies<sup>40</sup> | 8,293 | 1000 Genomes Phase 1 | Finnish | age, sex, BMI, population structure |
| sIL-6R levels | INTERVAL study<sup>41</sup> | 3,301 | 1000 Genomes Phase 3 | European | age, sex, duration between blood draw and processing, population structure |
| Fibrinogen levels | CHARGE Inflammation Working Group<sup>43</sup> | 120,246 | 1000 Genomes Phase 1 | European | age, sex, population structure |
| Ischemic stroke | MEGASTROKE Consortium<sup>32</sup> | 34,217/406,630 | 1000 Genomes Phase 1 | European | age, sex, population structure |
| Large artery stroke | MEGASTROKE Consortium<sup>32</sup> | 4,373/406,111 | 1000 Genomes Phase 1 | European | age, sex, population structure |
| Cardioembolic stroke | MEGASTROKE Consortium<sup>32</sup> | 7,193/406,111 | 1000 Genomes Phase 1 | European | age, sex, population structure |
| Small vessel stroke | MEGASTROKE Consortium<sup>32</sup> | 5,386/406,111 | 1000 Genomes Phase 1 | European | age, sex, population structure |
| Coronary artery disease | CARDIoGRAMplusC4D Consortium<sup>44</sup> | 60,801/123,504 | HapMap | European and Asian | age, sex, population structure |
| Myocardial infarction | CARDIoGRAMplusC4D Consortium<sup>44</sup> | 43,676/123,504 | HapMap | European and Asian | age, sex, population structure |
| Aortic aneurysm | UK Biobank<sup>81</sup> | 1,817/314,325 | HRC + UK10K | White British | age, sex, population structure, genotyping platform array |
| Carotid plaque | CHARGE Consortium<sup>82</sup> | 21,540/26894 | 1000 Genomes Phase 1 | European | age, sex, population structure |
| Peripheral artery disease | UK Biobank<sup>81</sup> | 3,992/313,725 | HRC + UK10K | White British | age, sex, population structure, genotyping platform array |
| Heart failure | UK Biobank<sup>81</sup> | 8,970/312,436 | HRC + UK10K | White British | age, sex, population structure, genotyping platform array |
| Atrial fibrillation | AFGen Consortium<sup>83</sup> | 18,398/111,433 | 1000 Genomes Phase 1 | European and Asian | age, sex, population structure |
| Venous thromboembolism | INVENT Consortium<sup>84</sup> | 7,507/52,632 | 1000 Genomes Phase 1 | European | age, sex, population structure |
| Deep vein thrombosis | UK Biobank<sup>81</sup> | 4,135/302,337 | HRC + UK10K | White British | age, sex, population structure, genotyping platform array |
| Pulmonary embolism | UK Biobank<sup>81</sup> | 5,400/302,186 | HRC + UK10K | White British | age, sex, population structure, genotyping platform array |

**Figure 1.**
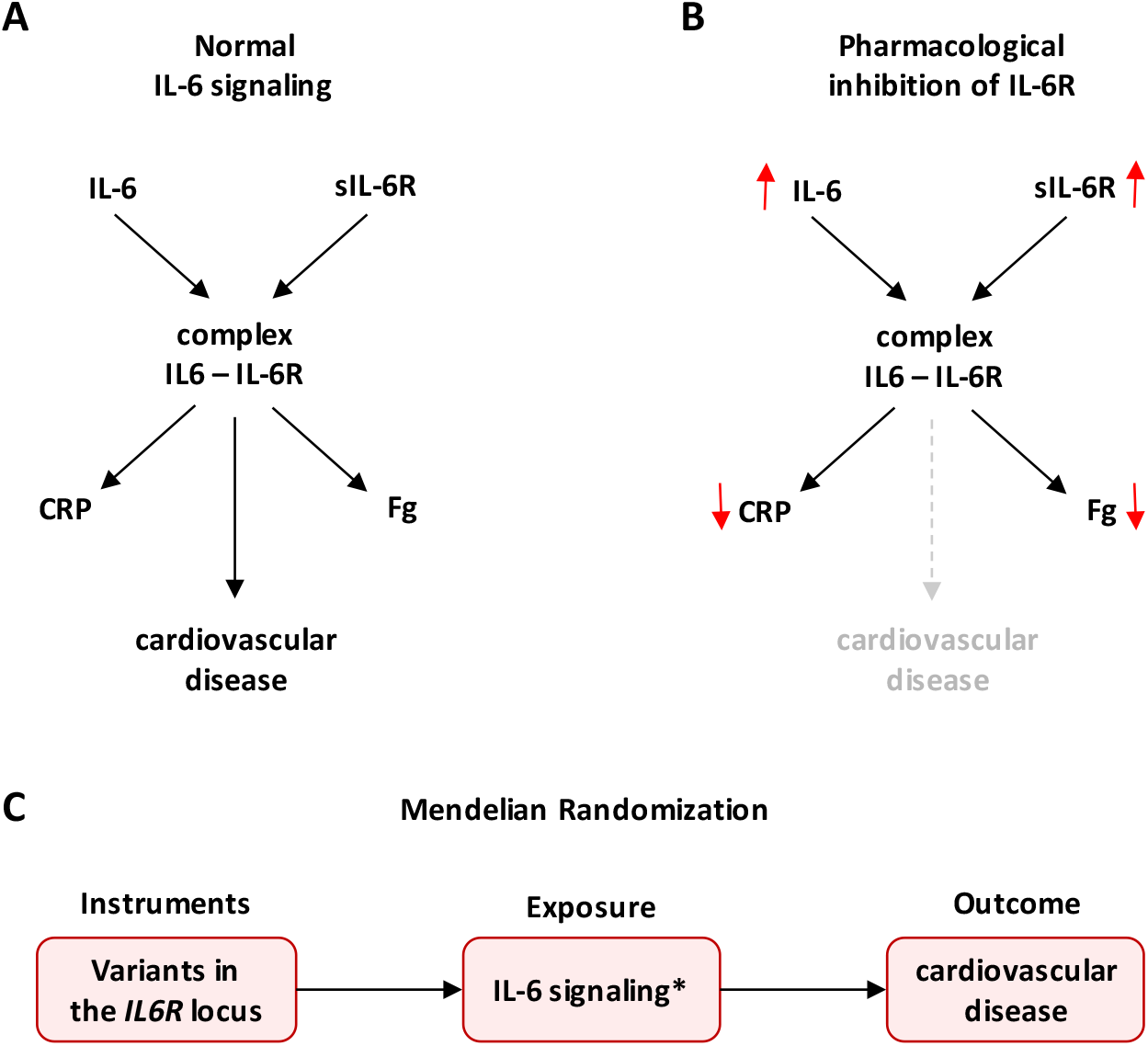
Conceptual framework and design of the current Mendelian Randomization approach. (**A**) Shown is a simplified scheme of IL-6 signaling, which is induced by binding of IL-6 to the soluble or the membrane-bound form of its receptor (IL-6R). IL-6 signaling results in increased C-reactive protein (CRP) and Fibrinogen (Fg) levels and is associated with a higher risk of cardiovascular disease. (**B**) Pharmacological inhibition of IL-6R leads to increases in the levels of upstream regulators (IL-6 and sIL-6R), and decreases in the levels of downstream effectors (CRP and Fg) of the IL-6 signaling pathway, but its effects on cardiovascular disease remain unknown. (**C**) In the current MR approach, we selected genetic variants within the *IL6R* locus, which significantly associated with lower CRP levels, as instruments (proxies) for a downregulated IL-6 signaling, and explored their effects on ischemic stroke, coronary artery disease and other cardiovascular disease phenotypes. *IL-6 signaling was determined by the effects of the instruments on CRP levels. The instruments were further validated by exploring their effects on other upstream regulators (IL-6, sIL-6-R) and downstream effectors (Fg) of IL-6 signaling. sIL-6R, soluble IL-6 receptor.

In sensitivity analyses, we restricted our selection of instruments to SNPs within the *IL6R* gene (GRCh37/hg19 coordinates: chr1:154,377,669-154,441,926), to avoid potential pleiotropic effects through genes neighboring *IL6R* and increase confidence in the effects of the instruments through IL-6 signaling. As the instruments used in the current setting were not identified based on established biological effects, but solely on the basis of their statistical associations with CRP levels, in an additional sensitivity analysis, we restricted our genetic instrument to a single SNP (rs2228145) within the *IL6R* gene with well-established biological effects leading to a downregulation of the IL-6 signaling.^20, 34, 38, 39^

To disentangle the effects of IL-6 signaling from the respective effects of CRP, we selected SNPs associated with CRP levels at genome-wide significance (*p*<5×10^−8^) throughout the genome and clumped them to *r*^*2*^ < 0.1. We then performed MR analyses using all these SNPs as instruments, and performed 10,000 permutations for each outcome using 7 randomly selected SNPs (the same number as those used as instruments for IL-6 signaling). We further performed MR analyses using SNPs at the *CRP* locus as instruments (within a region of 300 kB upstream or downstream to the *CRP* gene; GRCh37/hg19 coordinates: chr1: 159,382,079-159,984,379).

To validate the instruments, we explored their associations with circulating levels of IL-6 and soluble IL-6R, which have previously been reported to increase as a result of both pharmacological inhibition and genetic downregulation of IL-6 signaling.^20^ We further explored association with fibrinogen levels, which is a downstream effector of IL-6 signaling and decreases after its blockade.^20^ The effects of genetic variants on IL-6 levels were obtained from a GWAS of 8,293 healthy individuals of Finnish ancestry.^40^ For soluble IL-6R levels, we used the summary statistics from the INTERVAL study exploring the human plasma proteome,^41^ as made publicly available through the PhenoScanner database.^42^ For fibrinogen levels, we used GWAS data from the CHARGE Inflammation Working Group on 120,246 European individuals.^43^

### Outcomes

The primary outcomes for this study were ischemic stroke and coronary artery disease. Genetic association estimates for ischemic stroke and coronary artery disease were derived from the MEGASTROKE^32^ and CARDIoGRAMplusC4D^44^ consortia, respectively. Specifically, for ischemic stroke we used the European sub-dataset of MEGASTROKE (34,217 cases and 404,630 controls) to avoid population stratification with the CRP GWAS dataset, which also included solely individuals of European ancestry.^32^ The CARDIoGRAMplusC4D refers to a GWAS of 60,801 cases with coronary artery disease and 123,504 controls, primarily (77%) of European ancestry.^44^ Definitions for major ischemic stroke subtypes in MEGASTROKE followed the Trial of Org 10172 in Acute Stroke Treatment (TOAST) criteria with the following samples for analysis: large artery stroke (4,373 cases), cardioembolic stroke (7,193 cases), and small vessel stroke (5,386 cases; 404,630 controls for all subtypes).^45^ We further extended our analyses to other cardiovascular outcomes including myocardial infarction, aortic aneurysm, carotid artery plaque, peripheral artery disease, heart failure, atrial fibrillation, venous thromboembolism, deep vein thrombosis, and pulmonary embolism. The data sources and the sample sizes for these studies are presented in **Table 1**. For aortic aneurysm, heart failure, peripheral artery disease, deep vein thrombosis, and pulmonary embolism we used data from the UK Biobank, as described in **Supplementary Methods**.

### Mendelian Randomization analyses

After extracting the association estimates between the variants and the outcomes and harmonizing the direction of estimates by effect alleles, we computed MR estimates for each instrument with the Wald estimator and standard errors with the Delta method.^46^ We then pooled individual MR estimates using fixed-effects inverse-variance weighted (IVW) meta-analyses.^47^ To provide clinically relevant results, all effect estimates were scaled to the CRP-decreasing effect of tocilizumab (8 mg/kg), between 4 and 24 weeks after administration (a decrease of CRP levels by 67%), as determined by a meta-analysis of 4 clinical trials.^20^ For the main IVW analyses, we performed power calculations and estimated the minimum and maximum effects that we had 80% statistical power to detect.^48^

The IVW method was our primary MR analysis approach. Although the selection of instruments on a specific gene reduces the possibility of invalid variants,^49^ the derived estimates might still be biased in case of directional pleiotropy. Hence, we further applied sensitivity MR analyses that are more robust to the inclusion of pleiotropic variants: the weighted median estimator, the contamination mixture method, and the MR Pleiotropy Residual Sum and Outlier (MR-PRESSO). The weighted median estimator provides consistent estimates as long as at least half of the variants used in the MR analysis are valid.^50^ The contamination mixture method constructs a likelihood function of the individual estimates and under the assumption that the estimates of the valid instruments would follow a distribution centered around the causal effect and any invalid instruments would follow a distribution around zero, it calculates MR estimates that would maximize this likelihood.^51^ The contamination method assumes that only some of the genetic variants used are valid instruments and it has been found to perform better than other methods under the presence of invalid instruments.^52^ Finally, we applied MR-PRESSO, which regresses the SNP-outcome estimates against the SNP-exposure estimates to test, using residual errors, whether there are outlier SNPs. Outliers are detected by sequentially removing all genetic variants from the analyses and comparing the residual sum of squares as a global heterogeneity measure (p-value for detecting outliers <0.05).^53^ MR-PRESSO then removes the identified outliers and provides outlier-corrected MR estimates.^53^ MR-PRESSO, is outlier-robust, but still relies on the assumption that at least half of the variants are valid instruments.^53^

For the primary analyses (associations between downregulated IL-6 signaling and risk of ischemic stroke or coronary artery disease), we set a statistical significance threshold at a two-sided *p*-value of < 0.05. For ischemic stroke subtypes and for other cardiovascular outcomes, we corrected for multiple comparisons with the Bonferroni method. Thus, the statistical significance thresholds were set at *p*<0.05/3=0.017 for the 3 ischemic stroke subtypes, and at *p*<0.05/9=0.0055 for the 9 cardiovascular outcomes. Associations not reaching these thresholds, but showing *p*-values <0.05 were considered suggestive. All analyses were performed in R (v3.5.0; The R Foundation for Statistical Computing).

## RESULTS

### Identification and validation of genetic variants as proxies of downregulated IL-6 signaling

Using our pre-defined selection criteria, we identified 7 SNPs to serve as instruments for downregulated IL-6 signaling (**Table 2)**. Three of these instruments were situated within the *IL6R* gene (**Supplementary Figure 1**). The F-statistics of the 7 SNPs ranged from 81 to 764 indicating a low probability of weak instrument bias.^37^ Power calculations indicated that these instruments provide adequate statistical power (>80%) to detect ORs at the magnitude of 0.90 or lower for ischemic stroke and coronary artery disease regarding the effect of genetically downregulated IL-6 signaling (scaled to the CRP-decreasing effect of tocilizumab) (**Table S1**). We were further sufficiently powered (>80%) to detect ORs at the magnitude of 0.80 or lower for ischemic stroke subtypes.

**Table 2.** Single nucleotide polymorphisms (SNP) used in the current analyses for proxying the effects of IL-6 signaling. The betas, standard errors, and p-values refer to associations of these SNPs with CRP levels.

| SNP | Effect allele | Non-effect allele | Chromosome | MAF | Position (GRCh37/hg19) | Beta <sup>†</sup> | Standard Error | P-value | R<sup>2</sup> <sup>‡</sup> | F <sup>§</sup> |
| --- | --- | --- | --- | --- | --- | --- | --- | --- | --- | --- |
| rs73026617 | t | c | 1 | 0.097 | 154,369,981 | 0.0474 | 0.0068 | 3.16E-12 | 3.94E-04 | 80.5 |
| rs12083537* | a | g | 1 | 0.193 | 154,381,103 | 0.0643 | 0.0053 | 7.14E-34 | 1.29E-03 | 263.6 |
| rs4556348* | t | c | 1 | 0.148 | 154,394,296 | 0.0541 | 0.0067 | 6.77E-16 | 7.38E-04 | 151.0 |
| rs2228145* | a | c | 1 | 0.360 | 154,426,970 | 0.0899 | 0.0042 | 1.21E-101 | 3.72E-03 | 764.1 |
| rs11264224 | a | c | 1 | 0.193 | 154,568,086 | 0.0465 | 0.0057 | 3.41E-16 | 6.74E-04 | 137.8 |
| rs12059682 | t | c | 1 | 0.196 | 154,579,585 | -0.0441 | 0.0049 | 2.26E-19 | 6.13E-04 | 125.4 |
| rs34693607 | c | g | 1 | 0.184 | 154,661,369 | 0.0368 | 0.0057 | 1.07E-10 | 4.07E-04 | 83.2 |

\* Variants located within the *IL6R* gene.
<sup>†</sup> Beta coefficients correspond to 1-unit changes in the natural-log-transformed CRP (mg/L) per copy increment in effect allele.
<sup>‡</sup> $R^2 = 2 \times MAF \times (1 - MAF) \times beta^2$ , where MAF is the minimum allele frequency and beta is the effect estimate of the SNP on CRP levels.<sup>36</sup>
<sup>§</sup> $F = \frac{R^2 \times (N-2)}{1-R^2}$ where $R^2$ is the variance of CRP explained by the specific SNP (as explained above) and $N$ the number of individuals in the GWAS analysis.<sup>37</sup>

To validate the 7 instruments, we explored associations of genetically downregulated IL-6 signaling with circulating IL-6, soluble IL-6R, and fibrinogen levels. In accordance with randomized clinical trials testing the effects of tocilizumab versus placebo (8 mg/kg),^20^ genetically downregulated IL-6 signaling was associated with higher circulating IL-6 and soluble IL-6R levels and with lower circulating concentration of fibrinogen with the strongest effects seen for soluble IL-6R levels (**Figure 2**).

**Figure 2.**
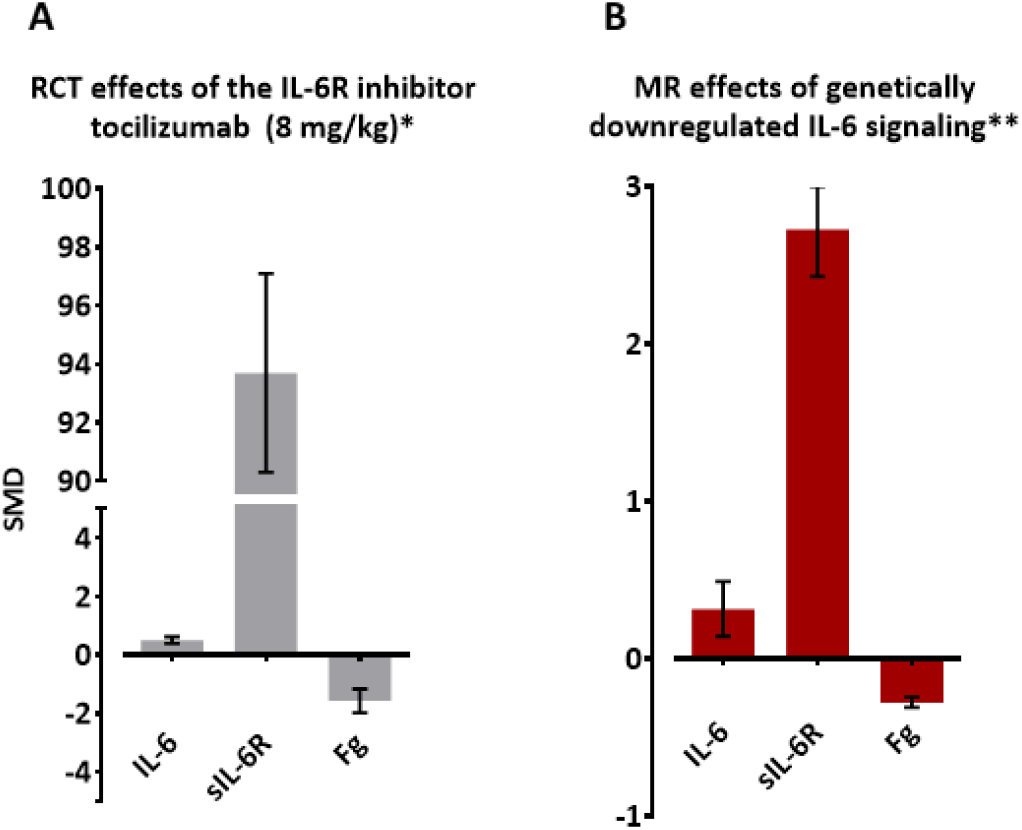
Effects of pharmacological inhibition of IL-6R and of genetic downregulation of IL-6 signaling on circulating levels of IL-6, soluble IL-6R (sIL-6R), and fibrinogen (Fg). **(A)** Effects of pharmacological inhibition of IL-6R on IL-6, sIL-6R, and Fg levels by administration of tocilizumab (8 mg/kg), as compared to placebo in a meta-analysis of 4 randomized clinical trials (RCT). Effects represent the standardized mean differences (SMD) in IL-6, sIL-6R, and Fg levels between 8 and 24 weeks after administration of tocilizumab (8 mg/kg), as compared to placebo. (**B**) Effects of genetic downregulation of IL-6 signaling on IL-6, sIL-6R, and Fg levels as determined by Mendelian Randomization (MR) analyses. Effects represent SMDs in IL-6, sIL-6R, and Fg levels. *The SMDs for RCTs are derived from a meta-analysis of 4 studies.^20^ ** The SMDs for the MR analyses are scaled to the CRP-decreasing effects of tocilizumab (8 mg/kg).

### Genetically downregulated IL-6 signaling, ischemic stroke and coronary artery disease

We next explored associations between genetically downregulated IL-6 signaling (scaled to the CRP-decreasing effect of tocilizumab) with the risk of ischemic stroke and coronary artery disease (**Figure 3**). In the primary IVW analysis, downregulated IL-6 signaling was associated with a lower risk of both ischemic stroke (OR: 0.89, 95%CI: 0.82-0.97, *p*=3⨯10^−3^) and coronary artery disease (OR: 0.84, 95%CI: 0.77-0.90, *p*=7⨯10^−6^). The alternative MR approaches (weighted median, contamination mixture, MR-PRESSO) all showed consistent association estimates (**Figure S2**).

**Figure 3.**
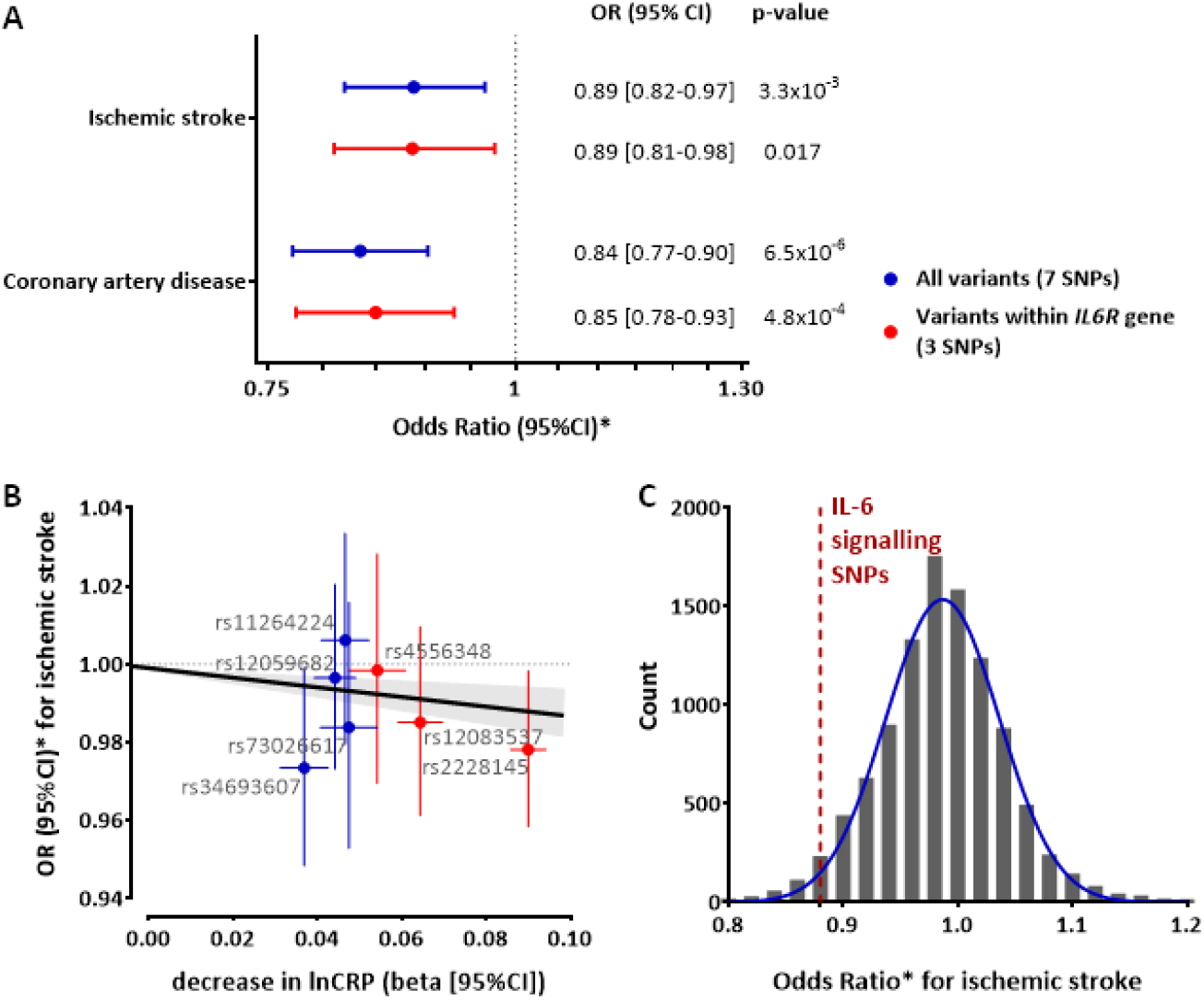
Mendelian Randomization associations of genetically downregulated IL-6 signaling with ischemic stroke. (**A**) Genetically downregulated IL-6 signaling in association with ischemic stroke and coronary artery disease as derived from IVW MR analyses either using the full set of genetic instruments (7 SNPs), or the restricted set of instruments (3 SNPs located within the *IL6R* gene). (**B**) SNP-specific effects regarding the associations of genetically downregulated IL-6 signaling with ischemic stroke and results derived from the IVW MR analysis. (**C**) Distributions of the effects of 7 randomly selected CRP-decreasing SNPs on risk of ischemic stroke and the position of the IL-6 signaling downregulating effect (7 SNPs included in our analyses). *Odds Ratios for genetically downregulated IL-6 signaling are scaled to the CRP-decreasing effects of tocilizumab (8 mg/kg).

In sensitivity analyses restricted to the 3 instruments within the *IL6R* gene, we likewise found genetically downregulated IL-6 signaling to be associated with a lower risk of ischemic stroke and coronary artery disease (**Figure 3** and **Figure S2**). Further restricting the analysis to a single SNP (rs2228145) with a well-described effect proxying the effects of pharmacological IL-6 signaling inhibition,^20^ we found presence of the allele linked to downregulated IL-6 signaling to be associated with lower risk of both ischemic stroke (OR: 0.88, 95%CI: 0.79-0.99, *p*=0.033) and coronary artery disease (OR: 0.75, 95%CI: 0.67-0.85, *p*=2⨯10^−7^).

To disentangle the effect of downregulated IL-6 signaling from the effect of CRP, we next performed MR analyses to explore associations between SNPs associated with CRP, and risk of ischemic stroke and coronary artery disease. These analyses showed no associations between genetically determined CRP levels and risk of either ischemic stroke or coronary artery disease independent of whether we used all variants reaching genome-wide significance (*p*<5⨯10^−8^) for association with CRP (187 SNPs), or whether we restricted the analyses to significant SNPs at the *CRP* locus (24 SNPs) (**Figure S3**). We further performed 10,000 permutations of MR analyses randomly selecting 7 out of the 187 SNPs associated with CRP. The effects of the 7 SNPs selected as instruments for downregulated IL-6 signaling on ischemic stroke and coronary artery disease were located at the 4^th^ and 1^st^ lowest percentiles of the respective distributions, corresponding to *p*-values of 0.04 and 0.01, respectively (**Figure 3C** and **Figure S4**), thus indicating that the effects of IL-6 signaling are independent of the effects of CRP itself.

### Genetically downregulated IL-6 signaling and ischemic stroke subtypes

Focusing on etiological stroke subtypes (**Figure 4**), we found genetic downregulation of IL-6 signaling to be associated with a lower risk of large artery stroke (OR: 0.76, 95%CI: 0.62-0.93, *p*=8⨯10^−3^) and small vessel stroke (OR: 0.71, 95%CI: 0.59-0.86, *p*=3⨯10^−4^), but not cardioembolic stroke (OR: 0.95, 95%CI: 0.74-1.22, *p*=0.667). The results were stable in all MR sensitivity analyses, including when restricting the analyses to the instruments within the *IL6R* gene (**Figure S5**). We further found no associations between genetically determined CRP levels, as determined by SNPs throughout the genome or SNPs at the *CRP* locus, and any of the ischemic stroke subtypes (**Figure S6**). Similarly, in permutations of analyses including 7 randomly allocated SNPs throughout the genome, the effects of the SNPs proxying the downregulated IL-6 signaling on large artery and small vessel stroke, were at the 3^rd^ and 0.1^th^ percentiles (corresponding to *p*-values of 0.03 and 0.001), respectively, thus supporting that the observed effects were again independent of CRP (**Figure S7**).

**Figure 4.**
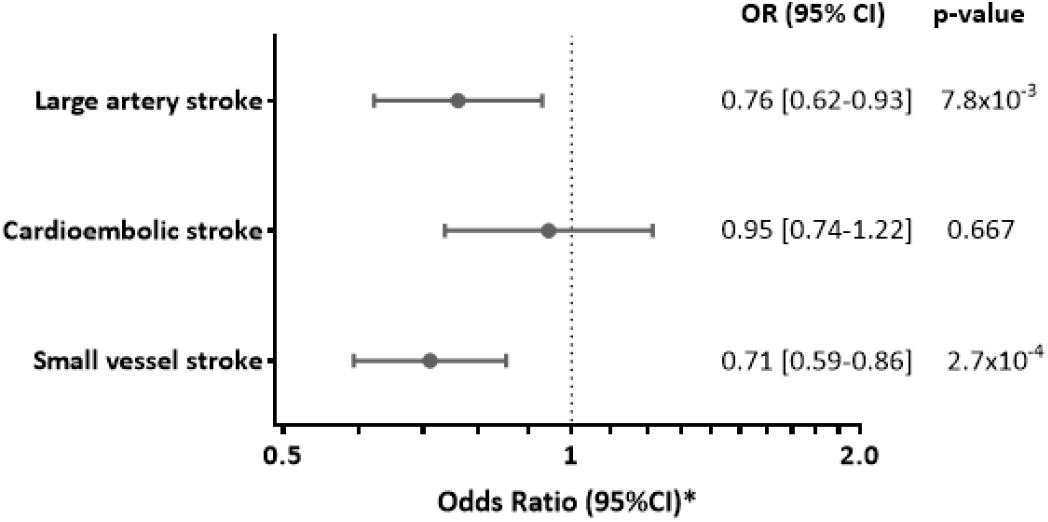
Mendelian Randomization associations of genetically downregulated IL-6 signaling with ischemic stroke etiological subtypes. The effects represent Odds Ratios (OR) derived from inverse-variance-weighted MR analyses. *Odds Ratios for genetically downregulated IL-6 signaling are scaled to the CRP-decreasing effects of tocilizumab (8 mg/kg).

### Genetically downregulated IL-6 signaling and other cardiovascular outcomes

In a last step, we expanded the analyses to other cardiovascular outcomes (**Figure 5**). Genetic predisposition to downregulated IL-6 signaling was associated with lower risks of myocardial infarction (OR: 0.88, 95%CI: 0.81-0.96, *p*=3⨯10^−3^) and aortic aneurysm (OR: 0.51, 95%CI: 0.37-0.68, *p*=1⨯10^−5^). We further found suggestive associations (*p*<0.05) with atrial fibrillation (OR: 0.83, 95%CI: 0.71-0.96, *p*=0.013) and carotid plaque (OR: 0.87, 95%CI: 0.77-0.99, *p*=0.041). In contrast, we found no significant associations with peripheral artery disease (OR: 0.91, 95%CI: 0.74-1.11, *p*=0.349), heart failure (OR: 0.90, 95%CI: 0.79-1.04, *p*=0.156), venous thromboembolism (OR: 0.98, 95%CI: 0.81-1.15, *p*=0.809), deep vein thrombosis (OR: 1.15, 95%CI: 0.94-1.40, *p*=0.183), and pulmonary embolism (OR: 0.92, 95%CI: 0.78-1.10, *p*=0.373).

**Figure 5.**
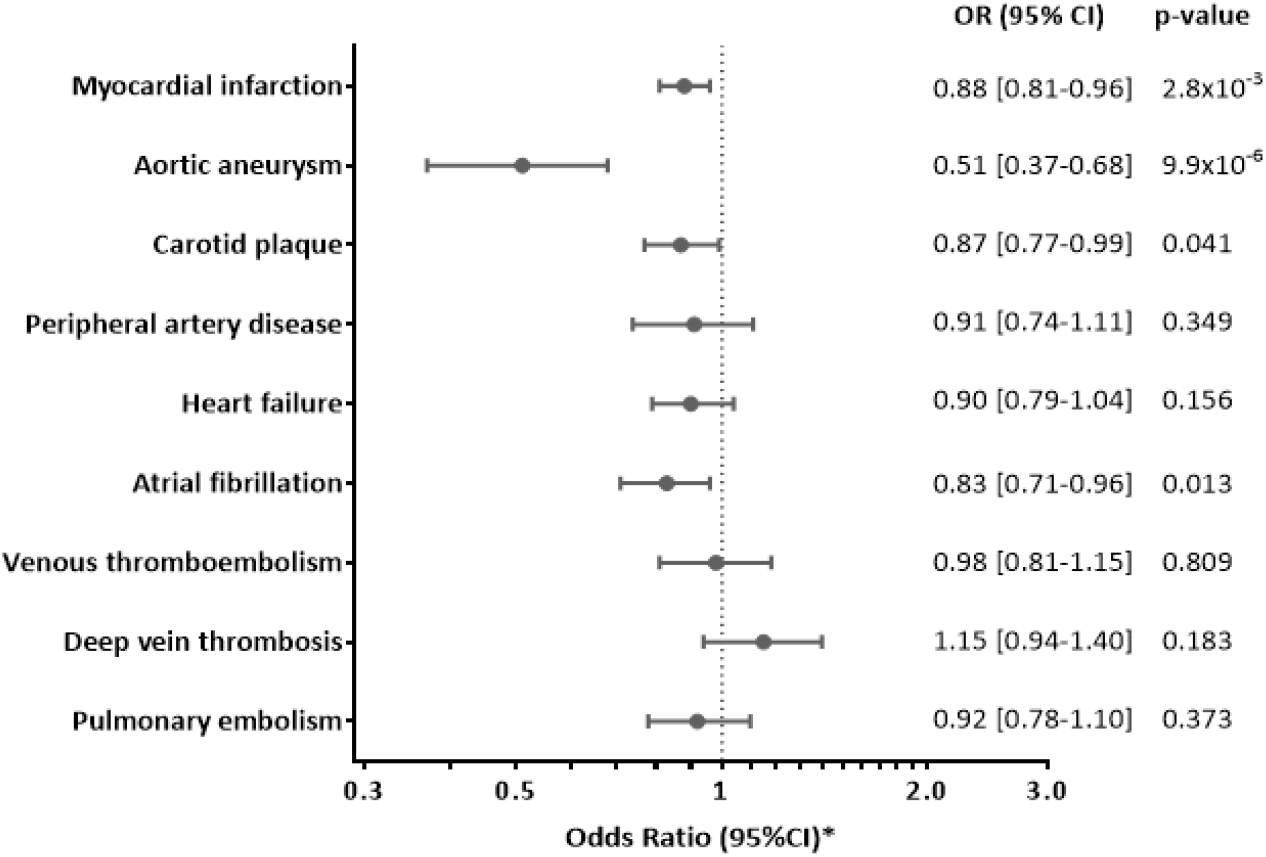
Mendelian Randomization associations of genetically downregulated IL-6 signaling with other cardiovascular outcomes. The effects represent Odds Ratios (OR) derived from inverse-variance-weighted MR analyses. *Odds Ratios for genetically downregulated IL-6 signaling are scaled to the CRP-decreasing effects of tocilizumab (8 mg/kg).

## DISCUSSION

Leveraging large-scale genetic data from multiple sources we identified variants serving as proxies for a genetic predisposition to downregulated IL-6 signaling and validated them using clinical trial data on pharmacological IL-6R inhibition. The identified proxies showed significant associations with a lower risk of both ischemic stroke and coronary artery disease. Among ischemic stroke subtypes, genetic predisposition to downregulated IL-6 signaling was associated with lower risks of large artery and small vessel stroke, but not cardioembolic stroke. Proxies for IL-6 signaling inhibition further showed significant associations with myocardial infarction and aortic aneurysm, and suggestive associations with atrial fibrillation and carotid plaque.

The MR association between genetically downregulated IL-6 signaling and lower risk of large artery stroke extends previous clinical,^17, 18, 21^ genetic,^19, 20^ and experimental^54, 55^ data demonstrating a key role of IL-6 signaling in atherosclerosis. By binding to IL-6R, IL-6 promotes downstream effects that include induction of macrophage recruitment^56^ and arterial smooth muscle cell proliferation,^55, 57^ and have been linked with plaque initiation,^58^ plaque destabilization,^54^ microvascular flow dysfunction,^59^ and adverse outcomes in the setting of acute ischemia.^60^ Moreover, pharmacological inhibition of IL-6R has been shown to attenuate atherosclerotic lesions in an experimental model of atherosclerosis.^61^ Our finding of an effect of genetic predisposition to downregulated IL-6 signaling on multiple atherosclerotic phenotypes (large artery stroke, coronary artery disease, myocardial infarction, aortic aneurysm, atrial fibrillation, carotid plaque) provides further support that IL-6 signaling is critically implicated in atherogenesis and atheroprogression and might represent a valid therapeutic target.

Notably, we found genetically downregulated IL-6 signaling to be further associated with small vessel stroke. There is only limited evidence regarding a role of inflammation in general and of IL-6 signaling in particular in cerebral small vessel disease.^62^ In a small prospective study of 123 patients with manifestations of cerebral small vessel disease, IL-6 circulating levels were associated with a higher risk of incident lacunes, a marker of small vessel disease on brain magnetic resonance imaging.^63^ However, cross-sectional analyses from larger population-based studies showed inconsistent findings for lacunes, silent brain infarcts and other manifestations of small vessel disease.^64-69^ While the specific mechanisms underlying our MR results remain unknown, our findings suggest that inhibition of IL-6 signaling aside from being a candidate treatment for atherosclerosis might also lower the risk of small vessel stroke.

Our results strongly support the candidacy of IL-6 signaling as a target for vascular prevention over and beyond previous data. The CANTOS trial targeted IL-1β rather than IL-6R and thus provided only indirect evidence for a benefit of interfering with IL-6 signaling.^11, 21^ Interestingly, the study further showed that part of the residual vascular risk after IL-1β inhibition could be explained by IL-6 levels, thus providing evidence that direct IL-6 signaling inhibition might represent a more effective strategy.^22^ Also, CANTOS was based on a population of individuals with coronary artery disease and explored a combined vascular endpoint rather than offering information on individual cardiovascular outcomes. With respect to stroke, there was a 7% reduction in incident stroke events in the IL-1β arm, which however did not reach statistical significance, possibly because of insufficient power.^8^ Our MR results provide evidence for directionally consistent effects of IL-6 signaling in multiple cardiovascular outcomes. Thus, our findings offer a solid basis for future clinical trials exploring the benefit of pharmacological IL-6R inhibition for the range of phenotypes examined here.

Interestingly, we found a particularly strong effect of genetically downregulated IL-6 signaling on aortic aneurysm. A role of IL-6 signaling in the pathogenesis of aortic aneurysm has been previously demonstrated by genetic studies.^38, 70, 71^ IL-6 signaling might contribute to the formation of aortic aneurysms through mechanisms aside from atherosclerosis, thus explaining the large effect. For instance, IL-6 signaling is a key pathway in the pathogenesis of large vessel vasculitides,^72^ which are strongly associated with the formation of aortic aneurysms.^16, 73, 74^

Our analysis provides no evidence for an association of genetically downregulated IL-6 signaling with cardioembolic stroke. In conjunction with the lack of significant MR associations with thrombotic phenotypes (venous thromboembolism, deep vein thrombosis, pulmonary embolism), our results do not support a role of IL-6 signaling in promoting coagulation and thrombosis. Yet, in accord with previous observational studies,^75-77^ we found IL-6 signaling to show a suggestive association with atrial fibrillation, the primary cause of cardioembolism and a common complication of coronary artery disease.^78, 79^ Given the relatively small magnitude of this association, any effect of IL-6 signaling on risk of cardioembolic stroke through atrial fibrillation would be expected to be small.

Our study has several strengths. Utilizing the most recent genetic data on CRP levels, ischemic stroke, and other cardiovascular phenotypes, we were sufficiently powered to show significant associations between genetically downregulated IL-6 signaling and multiple outcomes of interest. Using CRP levels, as a proxy for downstream IL-6 signaling enabled us to scale the derived association estimates to the respective effects of tocilizumab, as recorded in previous clinical trials, thus providing clinically meaningful estimates that might be comparable to those obtained from future trials. We further validated the effects of the selected proxies on upstream regulators (IL-6 and soluble IL-6R) and downstream effectors (fibrinogen) of IL-6 signaling, which were consistent with the effects observed with pharmacological inhibition of IL-6R. Finally, we could disentangle the effect of IL-6 signaling from the direct effect of CRP by determining the effects of CRP levels on risk of the examined outcomes and performing permutations for the effects of randomly selected CRP-decreasing variants.

Our study also has limitations. First, to proxy IL-6 signaling we used CRP levels, which are a downstream effect of the classical membrane-bound IL-6R-mediated signaling in hepatocytes.^80^ However, IL-6 also acts on other tissues not expressing the membrane-bound IL-6R, by binding to its soluble form, which is known as trans-signaling.^80^ Thus, our results may be interpreted as an effect of downstream regulation of classical IL-6 signaling but not IL-6 trans-signaling. Second, by design, our MR study assessed the effects of lifetime downregulated IL-6 signaling, which might differ from a shorter pharmacological inhibition. Third, there might be unknown pleiotropic effects of the genetic proxies used as instruments in the current study that might bias the associations. Of note, however, the results were remarkably consistent in sensitivity MR methods that are more robust to the inclusion of pleiotropic variants. Finally, our results were mainly based on individuals of European origin, and might thus not apply to other ethnic groups.

In conclusion, this study provides evidence for a causal effect of IL-6 signaling on ischemic stroke, particularly large artery and small vessel stroke, as well as a range of cardiovascular phenotypes. IL-6R blockade might represent a valid therapeutic target for lowering cardiovascular risk and should thus be further investigated in clinical trials.

## Data Availability

All analyses presented in the manuscript have been conducted with publically available data.
The instruments extracted for IL6 signaling are available in Table 1, for other authors to replicate our findings and use them in future work.

## Acknowledgements

We thank the following consortia for making data publicly available: MEGASTROKE Consortium, CARDIoGRAMplusC4D Consortium, AFGen Consortium, the YFS/FINRISK studies, and the INTERVAL study. This research has been conducted using the UK Biobank Resource (UK Biobank application 2532, “UK Biobank stroke study: developing an in-depth understanding of the determinants of stroke and its subtypes”).

## REFERENCES

1. Dalys GGBD and Collaborators H. Global, regional, and national disability-adjusted life-years (DALYs) for 315 diseases and injuries and healthy life expectancy (HALE), 1990-2015: a systematic analysis for the Global Burden of Disease Study 2015. Lancet. 2016;388:1603–1658.

2. Mortality GBD and Causes of Death C. Global, regional, and national life expectancy, all-cause mortality, and cause-specific mortality for 249 causes of death, 1980-2015: a systematic analysis for the Global Burden of Disease Study 2015. Lancet. 2016;388:1459–1544.

3. Collaborators GBDLRoS, Feigin VL, Nguyen G, Cercy K, Johnson CO, Alam T, Parmar PG, Abajobir AA, Abate KH, Abd-Allah F, Abejie AN, Abyu GY, Ademi Z, Agarwal G, Ahmed MB, Akinyemi RO, Al-Raddadi R, Aminde LN, Amlie-Lefond C, Ansari H, Asayesh H, Asgedom SW, Atey TM, Ayele HT, Banach M, Banerjee A, Barac A, Barker-Collo SL, Barnighausen T, Barregard L, Basu S, Bedi N, Behzadifar M, Bejot Y, Bennett DA, Bensenor IM, Berhe DF, Boneya DJ, Brainin M, Campos-Nonato IR, Caso V, Castaneda-Orjuela CA, Rivas JC, Catala-Lopez F, Christensen H, Criqui MH, Damasceno A, Dandona L, Dandona R, Davletov K, de Courten B, deVeber G, Dokova K, Edessa D, Endres M, Faraon EJA, Farvid MS, Fischer F, Foreman K, Forouzanfar MH, Gall SL, Gebrehiwot TT, Geleijnse JM, Gillum RF, Giroud M, Goulart AC, Gupta R, Gupta R, Hachinski V, Hamadeh RR, Hankey GJ, Hareri HA, Havmoeller R, Hay SI, Hegazy MI, Hibstu DT, James SL, Jeemon P, John D, Jonas JB, Jozwiak J, Kalani R, Kandel A, Kasaeian A, Kengne AP, Khader YS, Khan AR, Khang YH, Khubchandani J, Kim D, Kim YJ, Kivimaki M, Kokubo Y, Kolte D, Kopec JA, Kosen S, Kravchenko M, Krishnamurthi R, Kumar GA, Lafranconi A, Lavados PM, Legesse Y, Li Y, Liang X, Lo WD, Lorkowski S, Lotufo PA, Loy CT, Mackay MT, Abd El Razek HM, Mahdavi M, Majeed A, Malekzadeh R, Malta DC, Mamun AA, Mantovani LG, Martins SCO, Mate KK, Mazidi M, Mehata S, Meier T, Melaku YA, Mendoza W, Mensah GA, Meretoja A, Mezgebe HB, Miazgowski T, Miller TR, Ibrahim NM, Mohammed S, Mokdad AH, Moosazadeh M, Moran AE, Musa KI, Negoi RI, Nguyen M, Nguyen QL, Nguyen TH, Tran TT, Nguyen TT, Anggraini Ningrum DN, Norrving B, Noubiap JJ, O’Donnell MJ, Olagunju AT, Onuma OK, Owolabi MO, Parsaeian M, Patton GC, Piradov M, Pletcher MA, Pourmalek F, Prakash V, Qorbani M, Rahman M, Rahman MA, Rai RK, Ranta A, Rawaf D, Rawaf S, Renzaho AM, Robinson SR, Sahathevan R, Sahebkar A, Salomon JA, Santalucia P, Santos IS, Sartorius B, Schutte AE, Sepanlou SG, Shafieesabet A, Shaikh MA, Shamsizadeh M, Sheth KN, Sisay M, Shin MJ, Shiue I, Silva DAS, Sobngwi E, Soljak M, Sorensen RJD, Sposato LA, Stranges S, Suliankatchi RA, Tabares-Seisdedos R, Tanne D, Nguyen CT, Thakur JS, Thrift AG, Tirschwell DL, Topor-Madry R, Tran BX, Nguyen LT, Truelsen T, Tsilimparis N, Tyrovolas S, Ukwaja KN, Uthman OA, Varakin Y, Vasankari T, Venketasubramanian N, Vlassov VV, Wang W, Werdecker A, Wolfe CDA, Xu G, Yano Y, Yonemoto N, Yu C, Zaidi Z, El Sayed Zaki M, Zhou M, Ziaeian B, Zipkin B, Vos T, Naghavi M, Murray CJL and Roth GA. Global, Regional, and Country-Specific Lifetime Risks of Stroke, 1990 and 2016. N Engl J Med. 2018;379:2429–2437.

4. Feigin VL, Norrving B and Mensah GA. Global Burden of Stroke. Circ Res. 2017;120:439–448.

5. Esenwa CC and Elkind MS. Inflammatory risk factors, biomarkers and associated therapy in ischaemic stroke. Nat Rev Neurol. 2016;12:594–604.

6. Libby P, Ridker PM, Hansson GK and Leducq Transatlantic Network on Atherothrombosis. Inflammation in atherosclerosis: from pathophysiology to practice. J Am Coll Cardiol. 2009;54:2129–38.

7. Ridker PM. Targeting inflammatory pathways for the treatment of cardiovascular disease. Eur Heart J. 2014;35:540–3.

8. Ridker PM, Everett BM, Thuren T, MacFadyen JG, Chang WH, Ballantyne C, Fonseca F, Nicolau J, Koenig W, Anker SD, Kastelein JJP, Cornel JH, Pais P, Pella D, Genest J, Cifkova R, Lorenzatti A, Forster T, Kobalava Z, Vida-Simiti L, Flather M, Shimokawa H, Ogawa H, Dellborg M, Rossi PRF, Troquay RPT, Libby P, Glynn RJ and Group CT. Antiinflammatory Therapy with Canakinumab for Atherosclerotic Disease. N Engl J Med. 2017;377:1119–1131.

9. Georgakis MK, Gill D, Rannikmae K, Traylor M, Anderson CD, Lee JM, Kamatani Y, Hopewell JC, Worrall BB, Bernhagen J, Sudlow CLM, Malik R and Dichgans M. Genetically Determined Levels of Circulating Cytokines and Risk of Stroke. Circulation. 2019;139:256–268.

10. Georgakis MK, Malik R, Bjorkbacka H, Pana TA, Demissie S, Ayers C, Elhadad MA, Fornage M, Beiser AS, Benjamin EJ, Boekholdt MS, Engstrom G, Herder C, Hoogeveen RC, Koenig W, Melander O, Orho-Melander M, Schiopu A, Soderholm M, Wareham N, Ballantyne CM, Peters A, Seshadri S, Myint PK, Nilsson J, de Lemos JA and Dichgans M. Circulating Monocyte Chemoattractant Protein-1 and Risk of Stroke: A Meta-Analysis of Population-Based Studies Involving 17,180 Individuals. Circ Res. 2019.

11. Ridker PM. Anticytokine Agents: Targeting Interleukin Signaling Pathways for the Treatment of Atherothrombosis. Circ Res. 2019;124:437–450.

12. Scheller J, Chalaris A, Schmidt-Arras D and Rose-John S. The pro- and anti-inflammatory properties of the cytokine interleukin-6. Biochim Biophys Acta. 2011;1813:878–88.

13. Ridker PM. From C-Reactive Protein to Interleukin-6 to Interleukin-1: Moving Upstream To Identify Novel Targets for Atheroprotection. Circ Res. 2016;118:145–56.

14. Scott LJ. Tocilizumab: A Review in Rheumatoid Arthritis. Drugs. 2017;77:1865–1879.

15. Danese S, Vermeire S, Hellstern P, Panaccione R, Rogler G, Fraser G, Kohn A, Desreumaux P, Leong RW, Comer GM, Cataldi F, Banerjee A, Maguire MK, Li C, Rath N, Beebe J and Schreiber S. Randomised trial and open-label extension study of an anti-interleukin-6 antibody in Crohn’s disease (ANDANTE I and II). Gut. 2019;68:40–48.

16. Stone JH, Tuckwell K, Dimonaco S, Klearman M, Aringer M, Blockmans D, Brouwer E, Cid MC, Dasgupta B, Rech J, Salvarani C, Schett G, Schulze-Koops H, Spiera R, Unizony SH and Collinson N. Trial of Tocilizumab in Giant-Cell Arteritis. N Engl J Med. 2017;377:317–328.

17. Kaptoge S, Seshasai SR, Gao P, Freitag DF, Butterworth AS, Borglykke A, Di Angelantonio E, Gudnason V, Rumley A, Lowe GD, Jorgensen T and Danesh J. Inflammatory cytokines and risk of coronary heart disease: new prospective study and updated meta-analysis. Eur Heart J. 2014;35:578–89.

18. Ridker PM, Rifai N, Stampfer MJ and Hennekens CH. Plasma concentration of interleukin-6 and the risk of future myocardial infarction among apparently healthy men. Circulation. 2000;101:1767–72.

19. Collaboration IRGCERF, Sarwar N, Butterworth AS, Freitag DF, Gregson J, Willeit P, Gorman DN, Gao P, Saleheen D, Rendon A, Nelson CP, Braund PS, Hall AS, Chasman DI, Tybjaerg-Hansen A, Chambers JC, Benjamin EJ, Franks PW, Clarke R, Wilde AA, Trip MD, Steri M, Witteman JC, Qi L, van der Schoot CE, de Faire U, Erdmann J, Stringham HM, Koenig W, Rader DJ, Melzer D, Reich D, Psaty BM, Kleber ME, Panagiotakos DB, Willeit J, Wennberg P, Woodward M, Adamovic S, Rimm EB, Meade TW, Gillum RF, Shaffer JA, Hofman A, Onat A, Sundstrom J, Wassertheil-Smoller S, Mellstrom D, Gallacher J, Cushman M, Tracy RP, Kauhanen J, Karlsson M, Salonen JT, Wilhelmsen L, Amouyel P, Cantin B, Best LG, Ben-Shlomo Y, Manson JE, Davey-Smith G, de Bakker PI, O’Donnell CJ, Wilson JF, Wilson AG, Assimes TL, Jansson JO, Ohlsson C, Tivesten A, Ljunggren O, Reilly MP, Hamsten A, Ingelsson E, Cambien F, Hung J, Thomas GN, Boehnke M, Schunkert H, Asselbergs FW, Kastelein JJ, Gudnason V, Salomaa V, Harris TB, Kooner JS, Allin KH, Nordestgaard BG, Hopewell JC, Goodall AH, Ridker PM, Holm H, Watkins H, Ouwehand WH, Samani NJ, Kaptoge S, Di Angelantonio E, Harari O and Danesh J. Interleukin-6 receptor pathways in coronary heart disease: a collaborative meta-analysis of 82 studies. Lancet. 2012;379:1205–13.

20. Interleukin-6 Receptor Mendelian Randomisation Analysis C, Swerdlow DI, Holmes MV, Kuchenbaecker KB, Engmann JE, Shah T, Sofat R, Guo Y, Chung C, Peasey A, Pfister R, Mooijaart SP, Ireland HA, Leusink M, Langenberg C, Li KW, Palmen J, Howard P, Cooper JA, Drenos F, Hardy J, Nalls MA, Li YR, Lowe G, Stewart M, Bielinski SJ, Peto J, Timpson NJ, Gallacher J, Dunlop M, Houlston R, Tomlinson I, Tzoulaki I, Luan J, Boer JM, Forouhi NG, Onland-Moret NC, van der Schouw YT, Schnabel RB, Hubacek JA, Kubinova R, Baceviciene M, Tamosiunas A, Pajak A, Topor-Madry R, Malyutina S, Baldassarre D, Sennblad B, Tremoli E, de Faire U, Ferrucci L, Bandenelli S, Tanaka T, Meschia JF, Singleton A, Navis G, Mateo Leach I, Bakker SJ, Gansevoort RT, Ford I, Epstein SE, Burnett MS, Devaney JM, Jukema JW, Westendorp RG, Jan de Borst G, van der Graaf Y, de Jong PA, Mailand-van der Zee AH, Klungel OH, de Boer A, Doevendans PA, Stephens JW, Eaton CB, Robinson JG, Manson JE, Fowkes FG, Frayling TM, Price JF, Whincup PH, Morris RW, Lawlor DA, Smith GD, Ben-Shlomo Y, Redline S, Lange LA, Kumari M, Wareham NJ, Verschuren WM, Benjamin EJ, Whittaker JC, Hamsten A, Dudbridge F, Delaney JA, Wong A, Kuh D, Hardy R, Castillo BA, Connolly JJ, van der Harst P, Brunner EJ, Marmot MG, Wassel CL, Humphries SE, Talmud PJ, Kivimaki M, Asselbergs FW, Voevoda M, Bobak M, Pikhart H, Wilson JG, Hakonarson H, Reiner AP, Keating BJ, Sattar N, Hingorani AD and Casas JP. The interleukin-6 receptor as a target for prevention of coronary heart disease: a mendelian randomisation analysis. Lancet. 2012;379:1214–24.

21. Ridker PM, Libby P, MacFadyen JG, Thuren T, Ballantyne C, Fonseca F, Koenig W, Shimokawa H, Everett BM and Glynn RJ. Modulation of the interleukin-6 signalling pathway and incidence rates of atherosclerotic events and all-cause mortality: analyses from the Canakinumab Anti-Inflammatory Thrombosis Outcomes Study (CANTOS). Eur Heart J. 2018;39:3499–3507.

22. Ridker PM, MacFadyen JG, Thuren T and Libby P. Residual inflammatory risk associated with interleukin-18 and interleukin-6 after successful interleukin-1beta inhibition with canakinumab: further rationale for the development of targeted anti-cytokine therapies for the treatment of atherothrombosis. Eur Heart J. 2019.

23. Cainzos-Achirica M, Enjuanes C, Greenland P, McEvoy JW, Cushman M, Dardari Z, Nasir K, Budoff MJ, Al-Mallah MH, Yeboah J, Miedema MD, Blumenthal RS, Comin-Colet J and Blaha MJ. The prognostic value of interleukin 6 in multiple chronic diseases and all-cause death: The Multi-Ethnic Study of Atherosclerosis (MESA). Atherosclerosis. 2018;278:217–225.

24. Jenny NS, Callas PW, Judd SE, McClure LA, Kissela B, Zakai NA and Cushman M. Inflammatory cytokines and ischemic stroke risk: The REGARDS cohort. Neurology. 2019;92:e2375–e2384.

25. Smith GD and Ebrahim S. ‘Mendelian randomization’: can genetic epidemiology contribute to understanding environmental determinants of disease? Int J Epidemiol. 2003;32:1–22.

26. Kelly PJ, Murphy S, Coveney S, Purroy F, Lemmens R, Tsivgoulis G and Price C. Anti-inflammatory approaches to ischaemic stroke prevention. J Neurol Neurosurg Psychiatry. 2018;89:211–218.

27. Holmes MV, Ala-Korpela M and Smith GD. Mendelian randomization in cardiometabolic disease: challenges in evaluating causality. Nat Rev Cardiol. 2017;14:577–590.

28. Roberts R. Mendelian Randomization Studies Promise to Shorten the Journey to FDA Approval. JACC Basic Transl Sci. 2018;3:690–703.

29. Walker VM, Davey Smith G, Davies NM and Martin RM. Mendelian randomization: a novel approach for the prediction of adverse drug events and drug repurposing opportunities. Int J Epidemiol. 2017;46:2078–2089.

30. Gill D, Georgakis MK, Koskeridis F, Jiang L, Feng Q, Wei WQ, Theodoratou E, Elliott P, Denny JC, Malik R, Evangelou E, Dehghan A, Dichgans M and Tzoulaki I. Use of Genetic Variants Related to Antihypertensive Drugs to Inform on Efficacy and Side Effects. Circulation. 2019;140:270–279.

31. Ligthart S, Vaez A, Vosa U, Stathopoulou MG, de Vries PS, Prins BP, Van der Most PJ, Tanaka T, Naderi E, Rose LM, Wu Y, Karlsson R, Barbalic M, Lin H, Pool R, Zhu G, Mace A, Sidore C, Trompet S, Mangino M, Sabater-Lleal M, Kemp JP, Abbasi A, Kacprowski T, Verweij N, Smith AV, Huang T, Marzi C, Feitosa MF, Lohman KK, Kleber ME, Milaneschi Y, Mueller C, Huq M, Vlachopoulou E, Lyytikainen LP, Oldmeadow C, Deelen J, Perola M, Zhao JH, Feenstra B, LifeLines Cohort S, Amini M, Group CIW, Lahti J, Schraut KE, Fornage M, Suktitipat B, Chen WM, Li X, Nutile T, Malerba G, Luan J, Bak T, Schork N, Del Greco MF, Thiering E, Mahajan A, Marioni RE, Mihailov E, Eriksson J, Ozel AB, Zhang W, Nethander M, Cheng YC, Aslibekyan S, Ang W, Gandin I, Yengo L, Portas L, Kooperberg C, Hofer E, Rajan KB, Schurmann C, den Hollander W, Ahluwalia TS, Zhao J, Draisma HHM, Ford I, Timpson N, Teumer A, Huang H, Wahl S, Liu Y, Huang J, Uh HW, Geller F, Joshi PK, Yanek LR, Trabetti E, Lehne B, Vozzi D, Verbanck M, Biino G, Saba Y, Meulenbelt I, O’Connell JR, Laakso M, Giulianini F, Magnusson PKE, Ballantyne CM, Hottenga JJ, Montgomery GW, Rivadineira F, Rueedi R, Steri M, Herzig KH, Stott DJ, Menni C, Franberg M, St Pourcain B, Felix SB, Pers TH, Bakker SJL, Kraft P, Peters A, Vaidya D, Delgado G, Smit JH, Grossmann V, Sinisalo J, Seppala I, Williams SR, Holliday EG, Moed M, Langenberg C, Raikkonen K, Ding J, Campbell H, Sale MM, Chen YI, James AL, Ruggiero D, Soranzo N, Hartman CA, Smith EN, Berenson GS, Fuchsberger C, Hernandez D, Tiesler CMT, Giedraitis V, Liewald D, Fischer K, Mellstrom D, Larsson A, Wang Y, Scott WR, Lorentzon M, Beilby J, Ryan KA, Pennell CE, Vuckovic D, Balkau B, Concas MP, Schmidt R, Mendes de Leon CF, Bottinger EP, Kloppenburg M, Paternoster L, Boehnke M, Musk AW, Willemsen G, Evans DM, Madden PAF, Kahonen M, Kutalik Z, Zoledziewska M, Karhunen V, Kritchevsky SB, Sattar N, Lachance G, Clarke R, Harris TB, Raitakari OT, Attia JR, van Heemst D, Kajantie E, Sorice R, Gambaro G, Scott RA, Hicks AA, Ferrucci L, Standl M, Lindgren CM, Starr JM, Karlsson M, Lind L, Li JZ, Chambers JC, Mori TA, de Geus E, Heath AC, Martin NG, Auvinen J, Buckley BM, de Craen AJM, Waldenberger M, Strauch K, Meitinger T, Scott RJ, McEvoy M, Beekman M, Bombieri C, Ridker PM, Mohlke KL, Pedersen NL, Morrison AC, Boomsma DI, Whitfield JB, Strachan DP, Hofman A, Vollenweider P, Cucca F, Jarvelin MR, Jukema JW, Spector TD, Hamsten A, Zeller T, Uitterlinden AG, Nauck M, Gudnason V, Qi L, Grallert H, Borecki IB, Rotter JI, Marz W, Wild PS, Lokki ML, Boyle M, Salomaa V, Melbye M, Eriksson JG, Wilson JF, Penninx B, Becker DM, Worrall BB, Gibson G, Krauss RM, Ciullo M, Zaza G, Wareham NJ, Oldehinkel AJ, Palmer LJ, Murray SS, Pramstaller PP, Bandinelli S, Heinrich J, Ingelsson E, Deary IJ, Magi R, Vandenput L, van der Harst P, Desch KC, Kooner JS, Ohlsson C, Hayward C, Lehtimaki T, Shuldiner AR, Arnett DK, Beilin LJ, Robino A, Froguel P, Pirastu M, Jess T, Koenig W, Loos RJF, Evans DA, Schmidt H, Smith GD, Slagboom PE, Eiriksdottir G, Morris AP, Psaty BM, Tracy RP, Nolte IM, Boerwinkle E, Visvikis-Siest S, Reiner AP, Gross M, Bis JC, Franke L, Franco OH, Benjamin EJ, Chasman DI, Dupuis J, Snieder H, Dehghan A and Alizadeh BZ. Genome Analyses of >200,000 Individuals Identify 58 Loci for Chronic Inflammation and Highlight Pathways that Link Inflammation and Complex Disorders. Am J Hum Genet. 2018;103:691–706.

32. Malik R, Chauhan G, Traylor M, Sargurupremraj M, Okada Y, Mishra A, Rutten-Jacobs L, Giese AK, van der Laan SW, Gretarsdottir S, Anderson CD, Chong M, Adams HHH, Ago T, Almgren P, Amouyel P, Ay H, Bartz TM, Benavente OR, Bevan S, Boncoraglio GB, Brown RD, Jr., Butterworth AS, Carrera C, Carty CL, Chasman DI, Chen WM, Cole JW, Correa A, Cotlarciuc I, Cruchaga C, Danesh J, de Bakker PIW, DeStefano AL, den Hoed M, Duan Q, Engelter ST, Falcone GJ, Gottesman RF, Grewal RP, Gudnason V, Gustafsson S, Haessler J, Harris TB, Hassan A, Havulinna AS, Heckbert SR, Holliday EG, Howard G, Hsu FC, Hyacinth HI, Ikram MA, Ingelsson E, Irvin MR, Jian X, Jimenez-Conde J, Johnson JA, Jukema JW, Kanai M, Keene KL, Kissela BM, Kleindorfer DO, Kooperberg C, Kubo M, Lange LA, Langefeld CD, Langenberg C, Launer LJ, Lee JM, Lemmens R, Leys D, Lewis CM, Lin WY, Lindgren AG, Lorentzen E, Magnusson PK, Maguire J, Manichaikul A, McArdle PF, Meschia JF, Mitchell BD, Mosley TH, Nalls MA, Ninomiya T, O’Donnell MJ, Psaty BM, Pulit SL, Rannikmae K, Reiner AP, Rexrode KM, Rice K, Rich SS, Ridker PM, Rost NS, Rothwell PM, Rotter JI, Rundek T, Sacco RL, Sakaue S, Sale MM, Salomaa V, Sapkota BR, Schmidt R, Schmidt CO, Schminke U, Sharma P, Slowik A, Sudlow CLM, Tanislav C, Tatlisumak T, Taylor KD, Thijs VNS, Thorleifsson G, Thorsteinsdottir U, Tiedt S, Trompet S, Tzourio C, van Duijn CM, Walters M, Wareham NJ, Wassertheil-Smoller S, Wilson JG, Wiggins KL, Yang Q, Yusuf S, Bis JC, Pastinen T, Ruusalepp A, Schadt EE, Koplev S, Bjorkegren JLM, Codoni V, Civelek M, Smith NL, Tregouet DA, Christophersen IE, Roselli C, Lubitz SA, Ellinor PT, Tai ES, Kooner JS, Kato N, He J, van der Harst P, Elliott P, Chambers JC, Takeuchi F, Johnson AD, Sanghera DK, Melander O, Jern C, Strbian D, Fernandez-Cadenas I, Longstreth WT, Jr., Rolfs A, Hata J, Woo D, Rosand J, Pare G, Hopewell JC, Saleheen D, Stefansson K, Worrall BB, Kittner SJ, Seshadri S, Fornage M, Markus HS, Howson JMM, Kamatani Y, Debette S, Dichgans M, Malik R, Chauhan G, Traylor M, Sargurupremraj M, Okada Y, Mishra A, Rutten-Jacobs L, Giese AK, van der Laan SW, Gretarsdottir S, Anderson CD, Chong M, Adams HHH, Ago T, Almgren P, Amouyel P, Ay H, Bartz TM, Benavente OR, Bevan S, Boncoraglio GB, Brown RD, Jr., Butterworth AS, Carrera C, Carty CL, Chasman DI, Chen WM, Cole JW, Correa A, Cotlarciuc I, Cruchaga C, Danesh J, de Bakker PIW, DeStefano AL, Hoed MD, Duan Q, Engelter ST, Falcone GJ, Gottesman RF, Grewal RP, Gudnason V, Gustafsson S, Haessler J, Harris TB, Hassan A, Havulinna AS, Heckbert SR, Holliday EG, Howard G, Hsu FC, Hyacinth HI, Ikram MA, Ingelsson E, Irvin MR, Jian X, Jimenez-Conde J, Johnson JA, Jukema JW, Kanai M, Keene KL, Kissela BM, Kleindorfer DO, Kooperberg C, Kubo M, Lange LA, Langefeld CD, Langenberg C, Launer LJ, Lee JM, Lemmens R, Leys D, Lewis CM, Lin WY, Lindgren AG, Lorentzen E, Magnusson PK, Maguire J, Manichaikul A, McArdle PF, Meschia JF, Mitchell BD, Mosley TH, Nalls MA, Ninomiya T, O’Donnell MJ, Psaty BM, Pulit SL, Rannikmae K, Reiner AP, Rexrode KM, Rice K, Rich SS, Ridker PM, Rost NS, Rothwell PM, Rotter JI, Rundek T, Sacco RL, Sakaue S, Sale MM, Salomaa V, Sapkota BR, Schmidt R, Schmidt CO, Schminke U, Sharma P, Slowik A, Sudlow CLM, Tanislav C, Tatlisumak T, Taylor KD, Thijs VNS, Thorleifsson G, Thorsteinsdottir U, Tiedt S, Trompet S, Tzourio C, van Duijn CM, Walters M, Wareham NJ, Wassertheil-Smoller S, Wilson JG, Wiggins KL, Yang Q, Yusuf S, Amin N, Aparicio HS, Arnett DK, Attia J, Beiser AS, Berr C, Buring JE, Bustamante M, Caso V, Cheng YC, Choi SH, Chowhan A, Cullell N, Dartigues JF, Delavaran H, Delgado P, Dorr M, Engstrom G, Ford I, Gurpreet WS, Hamsten A, Heitsch L, Hozawa A, Ibanez L, Ilinca A, Ingelsson M, Iwasaki M, Jackson RD, Jood K, Jousilahti P, Kaffashian S, Kalra L, Kamouchi M, Kitazono T, Kjartansson O, Kloss M, Koudstaal PJ, Krupinski J, Labovitz DL, Laurie CC, Levi CR, Li L, Lind L, Lindgren CM, Lioutas V, Liu YM, Lopez OL, Makoto H, Martinez-Majander N, Matsuda K, Minegishi N, Montaner J, Morris AP, Muino E, Muller-Nurasyid M, Norrving B, Ogishima S, Parati EA, Peddareddygari LR, Pedersen NL, Pera J, Perola M, Pezzini A, Pileggi S, Rabionet R, Riba-Llena I, Ribases M, Romero JR, Roquer J, Rudd AG, Sarin AP, Sarju R, Sarnowski C, Sasaki M, Satizabal CL, Satoh M, Sattar N, Sawada N, Sibolt G, Sigurdsson A, Smith A, Sobue K, Soriano-Tarraga C, Stanne T, Stine OC, Stott DJ, Strauch K, Takai T, Tanaka H, Tanno K, Teumer A, Tomppo L, Torres-Aguila NP, Touze E, Tsugane S, Uitterlinden AG, Valdimarsson EM, van der Lee SJ, Volzke H, Wakai K, Weir D, Williams SR, Wolfe CDA, Wong Q, Xu H, Yamaji T, Sanghera DK, Melander O, Jern C, Strbian D, Fernandez-Cadenas I, Longstreth WT, Jr., Rolfs A, Hata J, Woo D, Rosand J, Pare G, Hopewell JC, Saleheen D, Stefansson K, Worrall BB, Kittner SJ, Seshadri S, Fornage M, Markus HS, Howson JMM, Kamatani Y, Debette S, Dichgans M, Consortium AF, Cohorts for H, Aging Research in Genomic Epidemiology C, International Genomics of Blood Pressure C, Consortium I, Starnet, BioBank Japan Cooperative Hospital G, Consortium C, Consortium E-C, Consortium EP-I, International Stroke Genetics C, Consortium M, Neurology Working Group of the CC, Network NSG, Study UKYLD, Consortium M and Consortium M. Multiancestry genome-wide association study of 520,000 subjects identifies 32 loci associated with stroke and stroke subtypes. Nat Genet. 2018;50:524–537.

33. Malik R, Rannikmae K, Traylor M, Georgakis MK, Sargurupremraj M, Markus HS, Hopewell JC, Debette S, Sudlow CLM, Dichgans M, consortium M and the International Stroke Genetics C. Genome-wide meta-analysis identifies 3 novel loci associated with stroke. Ann Neurol. 2018;84:934–939.

34. Parisinos CA, Serghiou S, Katsoulis M, George MJ, Patel RS, Hemingway H and Hingorani AD. Variation in Interleukin 6 Receptor Gene Associates With Risk of Crohn’s Disease and Ulcerative Colitis. Gastroenterology. 2018;155:303–306 e2.

35. Genomes Project C, Auton A, Brooks LD, Durbin RM, Garrison EP, Kang HM, Korbel JO, Marchini JL, McCarthy S, McVean GA and Abecasis GR. A global reference for human genetic variation. Nature. 2015;526:68–74.

36. Shim H, Chasman DI, Smith JD, Mora S, Ridker PM, Nickerson DA, Krauss RM and Stephens M. A multivariate genome-wide association analysis of 10 LDL subfractions, and their response to statin treatment, in 1868 Caucasians. PLoS One. 2015;10:e0120758.

37. Palmer TM, Lawlor DA, Harbord RM, Sheehan NA, Tobias JH, Timpson NJ, Davey Smith G and Sterne JA. Using multiple genetic variants as instrumental variables for modifiable risk factors. Stat Methods Med Res. 2012;21:223–42.

38. Cai T, Zhang Y, Ho YL, Link N, Sun J, Huang J, Cai TA, Damrauer S, Ahuja Y, Honerlaw J, Huang J, Costa L, Schubert P, Hong C, Gagnon D, Sun YV, Gaziano JM, Wilson P, Cho K, Tsao P, O’Donnell CJ, Liao KP and Program VAMV. Association of Interleukin 6 Receptor Variant With Cardiovascular Disease Effects of Interleukin 6 Receptor Blocking Therapy: A Phenome-Wide Association Study. JAMA Cardiol. 2018;3:849–857.

39. Ferreira RC, Freitag DF, Cutler AJ, Howson JM, Rainbow DB, Smyth DJ, Kaptoge S, Clarke P, Boreham C, Coulson RM, Pekalski ML, Chen WM, Onengut-Gumuscu S, Rich SS, Butterworth AS, Malarstig A, Danesh J and Todd JA. Functional IL6R 358Ala allele impairs classical IL-6 receptor signaling and influences risk of diverse inflammatory diseases. PLoS Genet. 2013;9:e1003444.

40. Ahola-Olli AV, Wurtz P, Havulinna AS, Aalto K, Pitkanen N, Lehtimaki T, Kahonen M, Lyytikainen LP, Raitoharju E, Seppala I, Sarin AP, Ripatti S, Palotie A, Perola M, Viikari JS, Jalkanen S, Maksimow M, Salomaa V, Salmi M, Kettunen J and Raitakari OT. Genome-wide Association Study Identifies 27 Loci Influencing Concentrations of Circulating Cytokines and Growth Factors. Am J Hum Genet. 2017;100:40–50.

41. Sun BB, Maranville JC, Peters JE, Stacey D, Staley JR, Blackshaw J, Burgess S, Jiang T, Paige E, Surendran P, Oliver-Williams C, Kamat MA, Prins BP, Wilcox SK, Zimmerman ES, Chi A, Bansal N, Spain SL, Wood AM, Morrell NW, Bradley JR, Janjic N, Roberts DJ, Ouwehand WH, Todd JA, Soranzo N, Suhre K, Paul DS, Fox CS, Plenge RM, Danesh J, Runz H and Butterworth AS. Genomic atlas of the human plasma proteome. Nature. 2018;558:73–79.

42. Kamat MA, Blackshaw JA, Young R, Surendran P, Burgess S, Danesh J, Butterworth AS and Staley JR. PhenoScanner V2: an expanded tool for searching human genotype-phenotype associations. Bioinformatics. 2019.

43. de Vries PS, Chasman DI, Sabater-Lleal M, Chen MH, Huffman JE, Steri M, Tang W, Teumer A, Marioni RE, Grossmann V, Hottenga JJ, Trompet S, Muller-Nurasyid M, Zhao JH, Brody JA, Kleber ME, Guo X, Wang JJ, Auer PL, Attia JR, Yanek LR, Ahluwalia TS, Lahti J, Venturini C, Tanaka T, Bielak LF, Joshi PK, Rocanin-Arjo A, Kolcic I, Navarro P, Rose LM, Oldmeadow C, Riess H, Mazur J, Basu S, Goel A, Yang Q, Ghanbari M, Willemsen G, Rumley A, Fiorillo E, de Craen AJ, Grotevendt A, Scott R, Taylor KD, Delgado GE, Yao J, Kifley A, Kooperberg C, Qayyum R, Lopez LM, Berentzen TL, Raikkonen K, Mangino M, Bandinelli S, Peyser PA, Wild S, Tregouet DA, Wright AF, Marten J, Zemunik T, Morrison AC, Sennblad B, Tofler G, de Maat MP, de Geus EJ, Lowe GD, Zoledziewska M, Sattar N, Binder H, Volker U, Waldenberger M, Khaw KT, McKnight B, Huang J, Jenny NS, Holliday EG, Qi L, McEvoy MG, Becker DM, Starr JM, Sarin AP, Hysi PG, Hernandez DG, Jhun MA, Campbell H, Hamsten A, Rivadeneira F, McArdle WL, Slagboom PE, Zeller T, Koenig W, Psaty BM, Haritunians T, Liu J, Palotie A, Uitterlinden AG, Stott DJ, Hofman A, Franco OH, Polasek O, Rudan I, Morange PE, Wilson JF, Kardia SL, Ferrucci L, Spector TD, Eriksson JG, Hansen T, Deary IJ, Becker LC, Scott RJ, Mitchell P, Marz W, Wareham NJ, Peters A, Greinacher A, Wild PS, Jukema JW, Boomsma DI, Hayward C, Cucca F, Tracy R, Watkins H, Reiner AP, Folsom AR, Ridker PM, O’Donnell CJ, Smith NL, Strachan DP and Dehghan A. A meta-analysis of 120 246 individuals identifies 18 new loci for fibrinogen concentration. Hum Mol Genet. 2016;25:358–70.

44. Nikpay M, Goel A, Won HH, Hall LM, Willenborg C, Kanoni S, Saleheen D, Kyriakou T, Nelson CP, Hopewell JC, Webb TR, Zeng L, Dehghan A, Alver M, Armasu SM, Auro K, Bjonnes A, Chasman DI, Chen S, Ford I, Franceschini N, Gieger C, Grace C, Gustafsson S, Huang J, Hwang SJ, Kim YK, Kleber ME, Lau KW, Lu X, Lu Y, Lyytikainen LP, Mihailov E, Morrison AC, Pervjakova N, Qu L, Rose LM, Salfati E, Saxena R, Scholz M, Smith AV, Tikkanen E, Uitterlinden A, Yang X, Zhang W, Zhao W, de Andrade M, de Vries PS, van Zuydam NR, Anand SS, Bertram L, Beutner F, Dedoussis G, Frossard P, Gauguier D, Goodall AH, Gottesman O, Haber M, Han BG, Huang J, Jalilzadeh S, Kessler T, Konig IR, Lannfelt L, Lieb W, Lind L, Lindgren CM, Lokki ML, Magnusson PK, Mallick NH, Mehra N, Meitinger T, Memon FU, Morris AP, Nieminen MS, Pedersen NL, Peters A, Rallidis LS, Rasheed A, Samuel M, Shah SH, Sinisalo J, Stirrups KE, Trompet S, Wang L, Zaman KS, Ardissino D, Boerwinkle E, Borecki IB, Bottinger EP, Buring JE, Chambers JC, Collins R, Cupples LA, Danesh J, Demuth I, Elosua R, Epstein SE, Esko T, Feitosa MF, Franco OH, Franzosi MG, Granger CB, Gu D, Gudnason V, Hall AS, Hamsten A, Harris TB, Hazen SL, Hengstenberg C, Hofman A, Ingelsson E, Iribarren C, Jukema JW, Karhunen PJ, Kim BJ, Kooner JS, Kullo IJ, Lehtimaki T, Loos RJF, Melander O, Metspalu A, Marz W, Palmer CN, Perola M, Quertermous T, Rader DJ, Ridker PM, Ripatti S, Roberts R, Salomaa V, Sanghera DK, Schwartz SM, Seedorf U, Stewart AF, Stott DJ, Thiery J, Zalloua PA, O’Donnell CJ, Reilly MP, Assimes TL, Thompson JR, Erdmann J, Clarke R, Watkins H, Kathiresan S, McPherson R, Deloukas P, Schunkert H, Samani NJ and Farrall M. A comprehensive 1,000 Genomes-based genome-wide association meta-analysis of coronary artery disease. Nat Genet. 2015;47:1121–1130.

45. Adams HP, Jr., Bendixen BH, Kappelle LJ, Biller J, Love BB, Gordon DL and Marsh EE, 3rd. Classification of subtype of acute ischemic stroke. Definitions for use in a multicenter clinical trial. TOAST. Trial of Org 10172 in Acute Stroke Treatment. Stroke. 1993;24:35–41.

46. Lawlor DA, Harbord RM, Sterne JA, Timpson N and Davey Smith G. Mendelian randomization: using genes as instruments for making causal inferences in epidemiology. Stat Med. 2008;27:1133–63.

47. Burgess S, Butterworth A and Thompson SG. Mendelian randomization analysis with multiple genetic variants using summarized data. Genet Epidemiol. 2013;37:658–65.

48. Brion MJ, Shakhbazov K and Visscher PM. Calculating statistical power in Mendelian randomization studies. Int J Epidemiol. 2013;42:1497–501.

49. Swerdlow DI, Kuchenbaecker KB, Shah S, Sofat R, Holmes MV, White J, Mindell JS, Kivimaki M, Brunner EJ, Whittaker JC, Casas JP and Hingorani AD. Selecting instruments for Mendelian randomization in the wake of genome-wide association studies. Int J Epidemiol. 2016;45:1600–1616.

50. Hartwig FP, Davey Smith G and Bowden J. Robust inference in summary data Mendelian randomization via the zero modal pleiotropy assumption. Int J Epidemiol. 2017;46:1985–1998.

51. Burgess S, Foley CN, Allara E, Staley JR and Howson JM. A robust and efficient method for Mendelian randomization with hundreds of genetic variants: unravelling mechanisms linking HDL-cholesterol and coronary heart disease. bioRxiv (Pre-print). 2019.

52. Slob EAW and Burgess S. A Comparison Of Robust Mendelian Randomization Methods Using Summary Data. bioRxiv (Pre-print). 2019.

53. Verbanck M, Chen CY, Neale B and Do R. Detection of widespread horizontal pleiotropy in causal relationships inferred from Mendelian randomization between complex traits and diseases. Nat Genet. 2018;50:693–698.

54. Huber SA, Sakkinen P, Conze D, Hardin N and Tracy R. Interleukin-6 exacerbates early atherosclerosis in mice. Arterioscler Thromb Vasc Biol. 1999;19:2364–7.

55. Ikeda U, Ikeda M, Oohara T, Oguchi A, Kamitani T, Tsuruya Y and Kano S. Interleukin 6 stimulates growth of vascular smooth muscle cells in a PDGF-dependent manner. Am J Physiol. 1991;260:H1713–7.

56. Jenkins BJ, Grail D, Inglese M, Quilici C, Bozinovski S, Wong P and Ernst M. Imbalanced gp130-dependent signaling in macrophages alters macrophage colony-stimulating factor responsiveness via regulation of c-fms expression. Mol Cell Biol. 2004;24:1453–63.

57. Wang D, Liu Z, Li Q, Karpurapu M, Kundumani-Sridharan V, Cao H, Dronadula N, Rizvi F, Bajpai AK, Zhang C, Muller-Newen G, Harris KW and Rao GN. An essential role for gp130 in neointima formation following arterial injury. Circ Res. 2007;100:807–16.

58. Schieffer B, Selle T, Hilfiker A, Hilfiker-Kleiner D, Grote K, Tietge UJ, Trautwein C, Luchtefeld M, Schmittkamp C, Heeneman S, Daemen MJ and Drexler H. Impact of interleukin-6 on plaque development and morphology in experimental atherosclerosis. Circulation. 2004;110:3493–500.

59. Guo F, Dong M, Ren F, Zhang C, Li J, Tao Z, Yang J and Li G. Association between local interleukin-6 levels and slow flow/microvascular dysfunction. J Thromb Thrombolysis. 2014;37:475–82.

60. Lindmark E, Diderholm E, Wallentin L and Siegbahn A. Relationship between interleukin 6 and mortality in patients with unstable coronary artery disease: effects of an early invasive or noninvasive strategy. JAMA. 2001;286:2107–13.

61. Akita K, Isoda K, Sato-Okabayashi Y, Kadoguchi T, Kitamura K, Ohtomo F, Shimada K and Daida H. An Interleukin-6 Receptor Antibody Suppresses Atherosclerosis in Atherogenic Mice. Front Cardiovasc Med. 2017;4:84.

62. Low A, Mak E, Rowe JB, Markus HS and O’Brien JT. Inflammation and cerebral small vessel disease: A systematic review. Ageing Res Rev. 2019;53:100916.

63. Staszewski J, Piusinska-Macoch R, Brodacki B, Skrobowska E and Stepien A. IL-6, PF-4, sCD40 L, and homocysteine are associated with the radiological progression of cerebral small-vessel disease: a 2-year follow-up study. Clin Interv Aging. 2018;13:1135–1141.

64. Shoamanesh A, Preis SR, Beiser AS, Vasan RS, Benjamin EJ, Kase CS, Wolf PA, DeCarli C, Romero JR and Seshadri S. Inflammatory biomarkers, cerebral microbleeds, and small vessel disease: Framingham Heart Study. Neurology. 2015;84:825–32.

65. Satizabal CL, Zhu YC, Mazoyer B, Dufouil C and Tzourio C. Circulating IL-6 and CRP are associated with MRI findings in the elderly: the 3C-Dijon Study. Neurology. 2012;78:720–7.

66. Hoshi T, Kitagawa K, Yamagami H, Furukado S, Hougaku H and Hori M. Relations of serum high-sensitivity C-reactive protein and interleukin-6 levels with silent brain infarction. Stroke. 2005;36:768–72.

67. Yoshida M, Tomitori H, Machi Y, Katagiri D, Ueda S, Horiguchi K, Kobayashi E, Saeki N, Nishimura K, Ishii I, Kashiwagi K and Igarashi K. Acrolein, IL-6 and CRP as markers of silent brain infarction. Atherosclerosis. 2009;203:557–62.

68. Fornage M, Chiang YA, O’Meara ES, Psaty BM, Reiner AP, Siscovick DS, Tracy RP and Longstreth WT, Jr. Biomarkers of Inflammation and MRI-Defined Small Vessel Disease of the Brain: The Cardiovascular Health Study. Stroke. 2008;39:1952–9.

69. Baune BT, Ponath G, Rothermundt M, Roesler A and Berger K. Association between cytokines and cerebral MRI changes in the aging brain. J Geriatr Psychiatry Neurol. 2009;22:23–34.

70. Jones GT, Tromp G, Kuivaniemi H, Gretarsdottir S, Baas AF, Giusti B, Strauss E, Van’t Hof FN, Webb TR, Erdman R, Ritchie MD, Elmore JR, Verma A, Pendergrass S, Kullo IJ, Ye Z, Peissig PL, Gottesman O, Verma SS, Malinowski J, Rasmussen-Torvik LJ, Borthwick KM, Smelser DT, Crosslin DR, de Andrade M, Ryer EJ, McCarty CA, Bottinger EP, Pacheco JA, Crawford DC, Carrell DS, Gerhard GS, Franklin DP, Carey DJ, Phillips VL, Williams MJ, Wei W, Blair R, Hill AA, Vasudevan TM, Lewis DR, Thomson IA, Krysa J, Hill GB, Roake J, Merriman TR, Oszkinis G, Galora S, Saracini C, Abbate R, Pulli R, Pratesi C, Saratzis A, Verissimo AR, Bumpstead S, Badger SA, Clough RE, Cockerill G, Hafez H, Scott DJ, Futers TS, Romaine SP, Bridge K, Griffin KJ, Bailey MA, Smith A, Thompson MM, van Bockxmeer FM, Matthiasson SE, Thorleifsson G, Thorsteinsdottir U, Blankensteijn JD, Teijink JA, Wijmenga C, de Graaf J, Kiemeney LA, Lindholt JS, Hughes A, Bradley DT, Stirrups K, Golledge J, Norman PE, Powell JT, Humphries SE, Hamby SE, Goodall AH, Nelson CP, Sakalihasan N, Courtois A, Ferrell RE, Eriksson P, Folkersen L, Franco-Cereceda A, Eicher JD, Johnson AD, Betsholtz C, Ruusalepp A, Franzen O, Schadt EE, Bjorkegren JL, Lipovich L, Drolet AM, Verhoeven EL, Zeebregts CJ, Geelkerken RH, van Sambeek MR, van Sterkenburg SM, de Vries JP, Stefansson K, Thompson JR, de Bakker PI, Deloukas P, Sayers RD, Harrison SC, van Rij AM, Samani NJ and Bown MJ. Meta-Analysis of Genome-Wide Association Studies for Abdominal Aortic Aneurysm Identifies Four New Disease-Specific Risk Loci. Circ Res. 2017;120:341–353.

71. Harrison SC, Smith AJ, Jones GT, Swerdlow DI, Rampuri R, Bown MJ, Aneurysm C, Folkersen L, Baas AF, de Borst GJ, Blankensteijn JD, Price JF, van der Graaf Y, McLachlan S, Agu O, Hofman A, Uitterlinden AG, Franco-Cereceda A, Ruigrok YM, Van’t Hof FN, Powell JT, van Rij AM, Casas JP, Eriksson P, Holmes MV, Asselbergs FW, Hingorani AD and Humphries SE. Interleukin-6 receptor pathways in abdominal aortic aneurysm. Eur Heart J. 2013;34:3707–16.

72. Terrades-Garcia N and Cid MC. Pathogenesis of giant-cell arteritis: how targeted therapies are influencing our understanding of the mechanisms involved. Rheumatology (Oxford). 2018;57:ii51–ii62.

73. Evans JM, Bowles CA, Bjornsson J, Mullany CJ and Hunder GG. Thoracic aortic aneurysm and rupture in giant cell arteritis. A descriptive study of 41 cases. Arthritis Rheum. 1994;37:1539–47.

74. Villiger PM, Adler S, Kuchen S, Wermelinger F, Dan D, Fiege V, Butikofer L, Seitz M and Reichenbach S. Tocilizumab for induction and maintenance of remission in giant cell arteritis: a phase 2, randomised, double-blind, placebo-controlled trial. Lancet. 2016;387:1921–7.

75. Marcus GM, Whooley MA, Glidden DV, Pawlikowska L, Zaroff JG and Olgin JE. Interleukin-6 and atrial fibrillation in patients with coronary artery disease: data from the Heart and Soul Study. Am Heart J. 2008;155:303–9.

76. Amdur RL, Mukherjee M, Go A, Barrows IR, Ramezani A, Shoji J, Reilly MP, Gnanaraj J, Deo R, Roas S, Keane M, Master S, Teal V, Soliman EZ, Yang P, Feldman H, Kusek JW, Tracy CM, Raj DS and Investigators CS. Interleukin-6 Is a Risk Factor for Atrial Fibrillation in Chronic Kidney Disease: Findings from the CRIC Study. PLoS One. 2016;11:e0148189.

77. Li J, Solus J, Chen Q, Rho YH, Milne G, Stein CM and Darbar D. Role of inflammation and oxidative stress in atrial fibrillation. Heart Rhythm. 2010;7:438–44.

78. Ntalla I, Kanoni S, Zeng L, Giannakopoulou O, Danesh J, Watkins H, Samani NJ, Deloukas P, Schunkert H and Group UKBCCCW. Genetic Risk Score for Coronary Disease Identifies Predispositions to Cardiovascular and Noncardiovascular Diseases. J Am Coll Cardiol. 2019;73:2932–2942.

79. Schmitt J, Duray G, Gersh BJ and Hohnloser SH. Atrial fibrillation in acute myocardial infarction: a systematic review of the incidence, clinical features and prognostic implications. Eur Heart J. 2009;30:1038–45.

80. Lissilaa R, Buatois V, Magistrelli G, Williams AS, Jones GW, Herren S, Shang L, Malinge P, Guilhot F, Chatel L, Hatterer E, Jones SA, Kosco-Vilbois MH and Ferlin WG. Although IL-6 trans-signaling is sufficient to drive local immune responses, classical IL-6 signaling is obligate for the induction of T cell-mediated autoimmunity. J Immunol. 2010;185:5512–21.

81. Sudlow C, Gallacher J, Allen N, Beral V, Burton P, Danesh J, Downey P, Elliott P, Green J, Landray M, Liu B, Matthews P, Ong G, Pell J, Silman A, Young A, Sprosen T, Peakman T and Collins R. UK biobank: an open access resource for identifying the causes of a wide range of complex diseases of middle and old age. PLoS Med. 2015;12:e1001779.

82. Franceschini N, Giambartolomei C, de Vries PS, Finan C, Bis JC, Huntley RP, Lovering RC, Tajuddin SM, Winkler TW, Graff M, Kavousi M, Dale C, Smith AV, Hofer E, van Leeuwen EM, Nolte IM, Lu L, Scholz M, Sargurupremraj M, Pitkanen N, Franzen O, Joshi PK, Noordam R, Marioni RE, Hwang SJ, Musani SK, Schminke U, Palmas W, Isaacs A, Correa A, Zonderman AB, Hofman A, Teumer A, Cox AJ, Uitterlinden AG, Wong A, Smit AJ, Newman AB, Britton A, Ruusalepp A, Sennblad B, Hedblad B, Pasaniuc B, Penninx BW, Langefeld CD, Wassel CL, Tzourio C, Fava C, Baldassarre D, O’Leary DH, Teupser D, Kuh D, Tremoli E, Mannarino E, Grossi E, Boerwinkle E, Schadt EE, Ingelsson E, Veglia F, Rivadeneira F, Beutner F, Chauhan G, Heiss G, Snieder H, Campbell H, Volzke H, Markus HS, Deary IJ, Jukema JW, de Graaf J, Price J, Pott J, Hopewell JC, Liang J, Thiery J, Engmann J, Gertow K, Rice K, Taylor KD, Dhana K, Kiemeney L, Lind L, Raffield LM, Launer LJ, Holdt LM, Dorr M, Dichgans M, Traylor M, Sitzer M, Kumari M, Kivimaki M, Nalls MA, Melander O, Raitakari O, Franco OH, Rueda-Ochoa OL, Roussos P, Whincup PH, Amouyel P, Giral P, Anugu P, Wong Q, Malik R, Rauramaa R, Burkhardt R, Hardy R, Schmidt R, de Mutsert R, Morris RW, Strawbridge RJ, Wannamethee SG, Hagg S, Shah S, McLachlan S, Trompet S, Seshadri S, Kurl S, Heckbert SR, Ring S, Harris TB, Lehtimaki T, Galesloot TE, Shah T, de Faire U, Plagnol V, Rosamond WD, Post W, Zhu X, Zhang X, Guo X, Saba Y, Consortium M, Dehghan A, Seldenrijk A, Morrison AC, Hamsten A, Psaty BM, van Duijn CM, Lawlor DA, Mook-Kanamori DO, Bowden DW, Schmidt H, Wilson JF, Wilson JG, Rotter JI, Wardlaw JM, Deanfield J, Halcox J, Lyytikainen LP, Loeffler M, Evans MK, Debette S, Humphries SE, Volker U, Gudnason V, Hingorani AD, Bjorkegren JLM, Casas JP and O’Donnell CJ. GWAS and colocalization analyses implicate carotid intima-media thickness and carotid plaque loci in cardiovascular outcomes. Nat Commun. 2018;9:5141.

83. Christophersen IE, Rienstra M, Roselli C, Yin X, Geelhoed B, Barnard J, Lin H, Arking DE, Smith AV, Albert CM, Chaffin M, Tucker NR, Li M, Klarin D, Bihlmeyer NA, Low SK, Weeke PE, Muller-Nurasyid M, Smith JG, Brody JA, Niemeijer MN, Dorr M, Trompet S, Huffman J, Gustafsson S, Schurmann C, Kleber ME, Lyytikainen LP, Seppala I, Malik R, Horimoto A, Perez M, Sinisalo J, Aeschbacher S, Theriault S, Yao J, Radmanesh F, Weiss S, Teumer A, Choi SH, Weng LC, Clauss S, Deo R, Rader DJ, Shah SH, Sun A, Hopewell JC, Debette S, Chauhan G, Yang Q, Worrall BB, Pare G, Kamatani Y, Hagemeijer YP, Verweij N, Siland JE, Kubo M, Smith JD, Van Wagoner DR, Bis JC, Perz S, Psaty BM, Ridker PM, Magnani JW, Harris TB, Launer LJ, Shoemaker MB, Padmanabhan S, Haessler J, Bartz TM, Waldenberger M, Lichtner P, Arendt M, Krieger JE, Kahonen M, Risch L, Mansur AJ, Peters A, Smith BH, Lind L, Scott SA, Lu Y, Bottinger EB, Hernesniemi J, Lindgren CM, Wong JA, Huang J, Eskola M, Morris AP, Ford I, Reiner AP, Delgado G, Chen LY, Chen YI, Sandhu RK, Li M, Boerwinkle E, Eisele L, Lannfelt L, Rost N, Anderson CD, Taylor KD, Campbell A, Magnusson PK, Porteous D, Hocking LJ, Vlachopoulou E, Pedersen NL, Nikus K, Orho-Melander M, Hamsten A, Heeringa J, Denny JC, Kriebel J, Darbar D, Newton-Cheh C, Shaffer C, Macfarlane PW, Heilmann-Heimbach S, Almgren P, Huang PL, Sotoodehnia N, Soliman EZ, Uitterlinden AG, Hofman A, Franco OH, Volker U, Jockel KH, Sinner MF, Lin HJ, Guo X, Isgc MCot, Neurology Working Group of the CC, Dichgans M, Ingelsson E, Kooperberg C, Melander O, Loos RJF, Laurikka J, Conen D, Rosand J, van der Harst P, Lokki ML, Kathiresan S, Pereira A, Jukema JW, Hayward C, Rotter JI, Marz W, Lehtimaki T, Stricker BH, Chung MK, Felix SB, Gudnason V, Alonso A, Roden DM, Kaab S, Chasman DI, Heckbert SR, Benjamin EJ, Tanaka T, Lunetta KL, Lubitz SA, Ellinor PT and Consortium AF. Large-scale analyses of common and rare variants identify 12 new loci associated with atrial fibrillation. Nat Genet. 2017;49:946–952.

84. Germain M, Chasman DI, de Haan H, Tang W, Lindstrom S, Weng LC, de Andrade M, de Visser MC, Wiggins KL, Suchon P, Saut N, Smadja DM, Le Gal G, van Hylckama Vlieg A, Di Narzo A, Hao K, Nelson CP, Rocanin-Arjo A, Folkersen L, Monajemi R, Rose LM, Brody JA, Slagboom E, Aissi D, Gagnon F, Deleuze JF, Deloukas P, Tzourio C, Dartigues JF, Berr C, Taylor KD, Civelek M, Eriksson P, Cardiogenics C, Psaty BM, Houwing-Duitermaat J, Goodall AH, Cambien F, Kraft P, Amouyel P, Samani NJ, Basu S, Ridker PM, Rosendaal FR, Kabrhel C, Folsom AR, Heit J, Reitsma PH, Tregouet DA, Smith NL and Morange PE. Meta-analysis of 65,734 individuals identifies TSPAN15 and SLC44A2 as two susceptibility loci for venous thromboembolism. Am J Hum Genet. 2015;96:532–42.

